# Artemisinin resistant *kelch13* R622I and RDT negativity approaching predominance in northern Ethiopia and emerging C580Y of African origin threaten falciparum malaria control

**DOI:** 10.1101/2025.06.23.25330019

**Authors:** Ayalew Jejaw Zeleke, Abebe A. Fola, George A. Tollefson, Karamako Niaré, Alec Leonetti, Om Taropawala, Jacob Marglous, Rebecca Crudale, Bokretsion G. Brhane, Ashenafi Assefa, Patience Kiyuka, Jonathan B. Parr, Asrat Hailu, Mulugeta Aemero, Jeffrey A. Bailey

## Abstract

The rise of antimalarial drug-resistant *Plasmodium falciparum* poses a major threat to malaria treatment, control, and elimination efforts. Mutations in the *kelch13* (*k13*) gene confer artemisinin partial resistance (ART-R), compromising the efficacy of combination therapies. In the Horn of Africa, the validated mutation *k13* R622I has rapidly emerged in parallel with other mutations elsewhere in Eastern Africa. To monitor and inform control efforts, we conducted in-depth sampling where R622I was first detected, using molecular inversion probe (MIP) targeted sequencing of key resistance mutations and informative SNPs across the genome. Samples were collected for a year within two districts, Gondar Zuria and Tach Armachiho, representing different ecological zones in northwest Ethiopia. Among 903 people with *P. falciparum* malaria, we observed a markedly higher prevalence of R622I (44.3%) compared to earlier reports, with significantly higher prevalence in Gondar Zuria (52%) than in Tach Armachiho (40%; p<0.001). Alarmingly, the prevalence of histidine rich protein 2 (HRP2) based rapid diagnostic test (RDT) negativity was higher in R622I mutants compared to wild types (48.3% vs 30.7%; p<0.001). Furthermore, 98.2% of R622I samples carried the multidrug resistance protein 1 NFD haplotype associated with reduced susceptibility to lumefantrine. We also detected the *k13* C580Y mutation in two patients (0.4%) from Gondar Zuria, the first report in the Horn of Africa. Identity-by-descent (IBD) analysis showed C580Y mutant parasites were likely clonal (IBD=1). Whole-genome sequencing confirmed their clonal nature and revealed flanking haplotypes distinct from those seen in Southeast Asia where the mutation was first observed, suggesting a local, de novo emergence. These findings highlight increasing prevalence and types of ART-R mutations and the concerning association now with RDT negativity that will further challenge malaria control and elimination efforts.

## Background

Malaria remains a major public health problem, especially in sub-Saharan Africa. In 2023, the global number of malaria cases and related deaths were 263 million and 597,000, representing a slight increase compared to 2022 (World malaria report 2024). Provision of effective treatment is critical to control malaria and progress towards elimination in addition to effective vector mitigation and control. Artemisinin-based combination therapies (ACTs) are the frontline treatment recommended by the World Health Organization (WHO) for uncomplicated *Plasmodium falciparum (Kunte and Kunwar 2011).* ACTs contain a fast-acting artemisinin derivative and a long-acting partner drug (Bosman and Mendis 2007) and have been adopted with the idea that they would slow the evolution of resistance (Nosten and White 2007). In Africa, ACTs were introduced in the early 2000s and have become the cornerstone of treatment across the continent, helping drive the reduction of malaria incidence and mortality (Nosten and White 2007; Slater et al. 2016). Artemether-lumefantrine (AL), Artesunate-amodiaquine (ASAQ), dihydroartemisinin-piperaquine (DP), artesunate-mefloquine (ASMQ), artesunate-sulfadoxine-pyrimethamine (ASSP), and artesunate-pyronaridine (AP) are approved ACTs in Africa. Among these, artemether-lumefantrine (AL) is the most common treatment for uncomplicated falciparum malaria in Africa accounting for 85% of all ACT purchases (Halsey and Plucinski 2023; Kokwaro, Mwai, and Nzila 2007).

Despite combination therapy, artemisinin partial resistance (ART-R) has emerged in Africa, posing a serious threat to malaria control efforts. ART-R is characterized by delayed parasite clearance within the first three days or a parasite clearance half-life (PCT_1/2_) of 5 hours or more after treatment with ACTs. ART-R is attributable to loss of function in the propeller domain of the *kelch13* (*k13*) gene, with various mutations in this domain leading to partial resistance. ART-R was first identified close to twenty years ago in Western Cambodia between 2006 and 2007 (Dondorp et al. 2009; Noedl et al. 2008), and quickly spread to other parts of the Greater Mekong (GM) subregion (Ashley et al. 2014). This spread was accompanied by the rapid emergence of partner drug resistance, leading to ACT clinical failure in Greater Mekong Subregion (GMS) (Parobek et al. 2017).

Recent studies have now identified both validated and candidate ART-R mutations in Africa. Since 2014, multiple mutations have been reported across countries including R561H in Rwanda (Uwimana et al. 2020), C469Y and A675V in Uganda (Conrad et al. 2023), and R622I in Eritrea (Mihreteab et al. 2023) and Ethiopia (Fola et al. 2023). *k13* P441L has recently emerged but lacks a known singular origin, suddenly appearing at sites in multiple countries: Tanzania (Connelly et al. 2025), Uganda (Conrad et al. 2023), Rwanda (Wernsman Young et al. 2025), Ethiopia (Brhane et al. 2024), Namibia (Eloff et al. 2025) and Zambia (Martin et al. 2025). Modeling of longitudinal data from Uganda supports that selection pressure favoring ART-R mutations is acting as strongly in Africa as it did in Southeast Asia (SEA) (Meier-Scherling et al. 2024). All flanking haplotypes for these mutations are distinct from SEA, consistent with de novo mutation in Africa rather than importation from SEA (Ishengoma et al. 2024; Connelly et al. 2025). Common mutations seen in SEA, C439A and C580Y, have not been observed in Africa--only fleeting reports in single samples without further confirmation. It remains unclear whether this absence reflects fundamental biological or selective differences between African and Asian parasite populations, or simply the stochastic emergence of different ART-R mutations across regions.

Three-fourths of Ethiopia’s land mass is malaria endemic, predominating in the west and with 60% of malaria cases caused by *P. falciparum* (Taffese et al. 2018). Transmission is highly heterogeneous and predominantly seasonal. Recently, malaria has resurged with cases increasing by 33% in 2022 compared to 2021 (from 1.3 to 3.3 million); and further to 4.1 million in 2023. Despite this resurgence, the country is still boldly committed to elimination by 2030. A key component of the elimination program is early diagnosis and treatment with effective medications (Bugssa and Tedla 2020). AL has been the first-line treatment for uncomplicated falciparum malaria since 2004 in the country (Lo et al. 2017). It remains efficacious (Abamecha et al. 2021), but detection of the ART-R *k13* R622I mutation (Bayih et al. 2016) and its observed rapid spread have raised concern (Fola et al. 2023). Research in the Amhara region of northwest Ethiopia, where the R622I was first identified in Africa (Bayih et al. 2016), has shown consistently increasing prevalence over time (Brhane et al. 2024). The emergence of *k13* mutations is compounded by additional challenges such as seasonal malaria transmission, population mobility, recent conflict, and the growing prevalence of *hrp2/3* gene deletions, which compromise the effectiveness of widely used rapid diagnostic tests (Feleke et al. 2021; Rogier et al. 2022; Gesesew et al. 2021). To investigate these converging threats, we conducted a genomic surveillance study using samples collected from 903 individuals with *Plasmodium falciparum* malaria over a one-year period in two districts of Ethiopia’s Amhara region: Gondar Zuria, a highland area with moderate transmission, and Tach Armachiho, a lowland district with high transmission intensity.

## Results

### Study population

A total of 903 individuals with confirmed *P. falciparum* infections by microscopy were enrolled into the study. Out of these, 55.3% (n=500) and 44.6% (n=403) originated from Gondar Zuria and Tach Armachiho districts, respectively (**Figure 1A**) across 26 villages called Kebeles. Of all participants, 59.4% were male (536/903), and the mean age was 22.1±13.0 years. 56 (6.2%) reporting past treatment and 65 participants (7%) reporting recent travel. The median parasite density of these samples using microscopy was 9121 parasites/μL (IQR 2519, 22160) (**Figure S1**), respectively. Microscopy detected gametocytemia was found in only 39 (4.4%) samples (**Table 1**).

**Figure 1:**
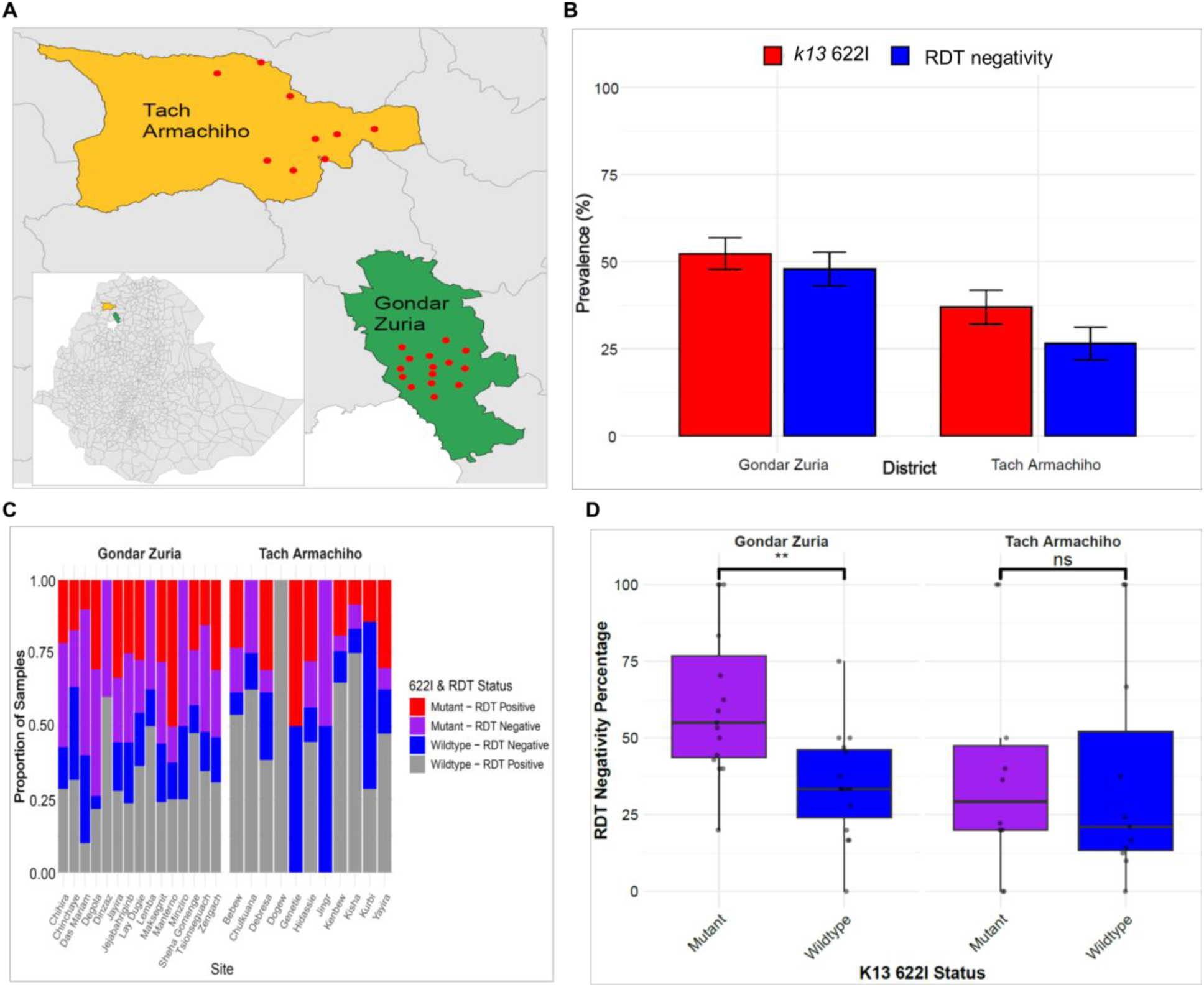
Prevalences of *k13* 622I mutation and RDT negativity in *P. falciparum* infections in Northwest Ethiopia. **A**) Map of study districts and Kabeles (villages - red dots) where participants were sampled. **B**) Grouped bar plot showing *k13* 622I prevalence and HRP2-based RDT negativity rate by district (all enrolled participants had microscopically confirmed *P. falciparum* infection). **C**) Stacked bar charts showing the distribution of 622I mutation by RDT result by Kebele. **D**) Prevalence of HRP2-based RDT negativity by *k13* 622I status and district showing significantly higher RDT negativity in mutant infections compared to wildtype infections in Gondar Zuria (Student’s t-test p=0.0022). No significant difference in RDT negativity percentage was found in Tach Armachiho. Boxplot center lines show the median value, with upper and lower bounds representing the 25th and 75th quartiles, and whiskers extending to 1.5×IQR from the lower and upper quartiles. Asterisks (*) denote level of statistically significant differences based on Student’s t-test: p < 0.01 (**), and p ≥ 0.05 (ns, not significant).

**Table 1:** Socio-demographic and clinical characteristics of study participants.

| Variables |  | Gondar Zuria<br>(n=500) | Tach Armachiho<br>(n=403) | Overall<br>(N=903) |
| --- | --- | --- | --- | --- |
| Sex | Male | 333(66.6%) | 203(50.4%) | 536 (59.4%) |
|  | Female | 167(33.4%) | 200(49.6%) | 367 (40.6%) |
| Age in years | <5 | 3(0.6%) | 7(1.7%) | 10 (1.1%) |
|  | 5-17 | 142(28.4) | 183(45.4%) | 359 (39.8%) |
|  | ≥18 | 355(71.0%) | 213(52.9%) | 534 (59.1%) |
| Residence | Rural | 382(76.4%) | 176(43.7%) | 558 (61.8%) |
|  | Urban | 118(23.6%) | 227(56.3%) | 345 (38.2%) |
| HRP2-based RDT | Positive | 230(46.0%) | 260(64.5%) | 490 (54.3%) |
|  | Negative | 218(43.6%) | 99(24.6%) | 317 (35.1%) |
|  | Not done | 52(10.4%) | 44(10.9%) | 96(10.6%) |
| Treatment history | Yes | 36(7.2%) | 20(5.0%) | 56 (6.2%) |
|  | No | 464(92.8%) | 383(95.0%) | 847 (92.8%) |
| Travel history | Yes | 44(8.8%) | 21(5.2%) | 65 (7.2%) |
|  | No | 456(91.2%) | 382(94.8%) | 838 (92.9%) |
| Gametocyte | Positive | 19(3.8%) | 20(5.0%) | 39 (4.4%) |
|  | Negative | 481(96.2%) | 383(95.0%) | 864 (95.6%) |
| Successfully<br>sequenced samples<br>for each target gene | *k13* | 486(97.2%) | 382(94.8%) | 868 (96.1%) |
|  | *mdr1* | 465(93%) | 365(90.6%) | 830 (91.9%) |
|  | *crt* | 438(87.6%) | 361(89.6%) | 799(88.5%) |
|  | *dhfr* | 451(90.2%) | 360(89.3%) | 811 (89.8%) |
|  | *dhps* | 450(90%) | 357(88.6) | 807 (89.3%) |

Of the 807 individuals with HRP2-based RDT results, 39.3% (317/807) tested negative despite being positive by light microscopy, indicating a substantial false-negative rate and raising concerns about the reliability of HRP2-based RDTs for detecting *P. falciparum* in this setting (**Table 1**). The mean parasite density in RDT-negative cases (GM=6388, 95% CI 5172-7889) was not significantly different from RDT-positive cases (GM=7862 95% CI 6778-9119) (Kruskal–Wallis rank test, p=0.1396). This suggests that RDT-negativity is unlikely due to low parasite density and is more likely attributable to *hrp2*/3 gene deletions, as previously reported (Feleke et al. 2021). The RDT-negativity rate was significantly higher in Gondar Zuria (48.7%) than in Tach Armachiho district (27.6%), (Pearson *χ*2 = 37.1451 p< 0.001, **Figure 1B**).

### Prevalence and local spread of ART-R mutations and its co-occurrence with RDT negativity

Using MIP targeted sequencing, we assessed the entire *k13* gene for prevalence of WHO-validated and candidate mutations as well as any other variations. The validated *k13* R622I mutation was found in 369/833 samples (44.3%, 95% CI 40.9-47.7%) (**Figure S2** and **Table S1**). For this mutation, 83.7% (309/369) of infections were solely mutant, while the remaining 16.3% (60/369) were mixed infections with wildtype. The prevalence of R622I was significantly (Student’s t-test p = 8.95e-6) higher in Gondar Zuria district (highland with lower malaria transmission) (51.4%, 95% CI 46.9-56.0%) compared to Tach Armachiho (lowland with higher malaria transmission) (35.4%, 95% CI 30.5-40.3%) (**Figure 1B** and **Table S2**). The mutation was identified in all but one Kebele (village) (**Figure 1C** and **Table S3)** across both districts suggesting broad spread of this mutation at local scale. There was no significant variation by Kebele (**Figure S3**). We also detected C580Y, a previously validated marker in SEA, in two individuals (0.2%). Other observed variations were outside of the propeller domain, with only the known common polymorphism K189T (25.2%), E401Q (3.5%) rising above 1% (**Table S1**).

Similar to R622I, RDT-negative and combined R622I RDT-negative parasites were prevalent in nearly all Kebeles across both districts (**Figure 1C**). RDT-negativity of 48.3% (95% CI 43.0-53.5%) for parasites with the 622I mutation was significantly higher than the 30.7% (95% CI 26.3-35.1%) observed in parasites without the mutation (Student’s t-test p<0.05, **Figure S4**). A similar pattern was observed at the district level. In Gondar Zuria, the RDT negativity rate was significantly higher among mutants (56.2%) compared to wildtypes (39.8%, Student’s t-test p = 0.0022). Likewise, in Tach Armachiho, the RDT negativity rate was higher among mutants (34.7%) compared to wildtypes (21.9%) but not statistically significant by Student’s t-test (**Figure 1D**).

### Emergence of *k13* C580Y mutation in Ethiopia

We detected two cases (0.2%) of the *k13* C580Y mutation, a well-established artemisinin resistance marker common in SEA but previously reported only once in Africa from Ghana (Aninagyei et al. 2020) and Nigeria (Martín Ramírez et al. 2025) without confirmatory analysis. MIP sequencing performed to examine their origin showed the two samples from Gondar Zuria district and Jejabahiriginb Kebele to be monogenomic, clonal infections (IBD=1). We further conducted whole-genome sequencing (WGS) using long-read Oxford Nanopore Technology (ONT) following selective whole-genome amplification (sWGA), confirming both were C580Y with no wildtype reads. We achieved sufficient coverage for flanking haplotype analysis in one sample, with an average genome coverage of 51X, and 82% of the genome covered at a depth of at least 5X (**Figure S5A**). The other sample has validated C580Y but had insufficient coverage (mean coverage 6X, and 36% of the genome had at least 5X coverage) (**Figure S5B**). The flanking haplotype based on the long-reads from the more deeply sequenced sample was distinct from those in SEA based on comparison to 41 representative publicly-available SEA C580Y parasites (**Figure 2**) (MalariaGEN et al. 2023). Interestingly, both C580Y mutant infections were HRP2-based RDT negative, despite having high parasitemia levels of 28,627 and 12,277 parasites/µL. Whole genome sequencing (WGS) analysis confirmed the absence of reads mapping to both the *hrp2* and *hrp3* genes in both samples (**FigureS 5C** and **5D**).

**Figure 2.**
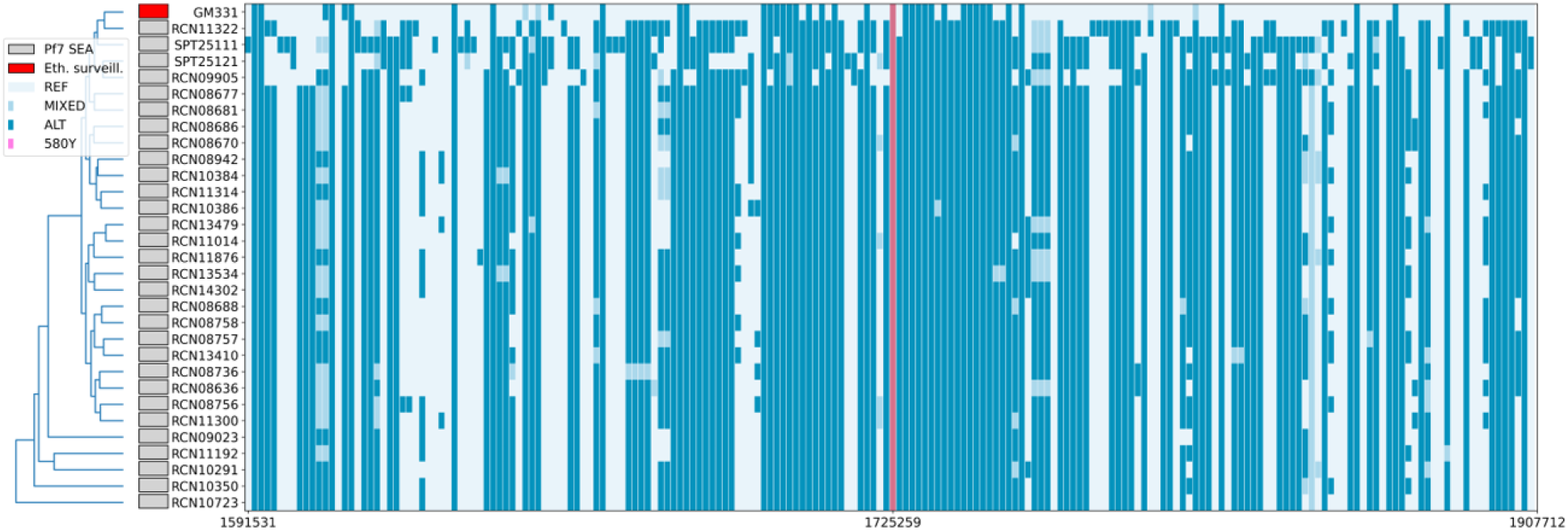
Flanking haplotype comparison of the *k13* locus in Ethiopian C580Y WGS sample compared to SEA *k13* C580Y mutants from Pf7 public database. Dark blue = alternate allele, light blue = reference allele (relative to 3D7) and intermediate blue = mixed genotype.

### Prevalence of other key antimalarial drug resistance markers

In addition to *k13* mutations, we observed near fixation of the MDR1 NFD haplotype (N86, 184F, D1246), associated with reduced lumefantrine susceptibility. Wild-type N86 and D1246 were found in 99.8% and 100% of samples, respectively, while 184F was present in 96.8%. Other *mdr1* mutations (e.g., A989E, R627I, D650N, T172K) were rare (<1%) (**Table S1**). For chloroquine resistance transporter (*crt*) gene, the resistant CVIET haplotype was found in 98.1% of samples, with co-occurring A220S (84.9%), N326S (76.4%), and R371I (92.5%). Unlike other East African regions where wild-type alleles are re-emerging, resistant haplotypes persist in Ethiopia, likely driven by continued chloroquine use for *P. vivax (Millet et al. 2004)*) (**Figure S6A**).

In Ethiopia, as in other African countries, SP was discontinued as a frontline antimalarial in 2004. It is also not used for intermittent preventive treatment for pregnant women (IPTp). SP resistance mutations remained common despite discontinued use. Mutations in the *Plasmodium falciparum* dihydrofolate reductase (*dhfr*) gene, N51I and S108N, were detected in 99.4% and 99.2% of samples, respectively, while C59R was present in 31.1% and I164L was absent. In the dihydropteroate synthase (*dhps*) gene, A437G and K540E mutations were found in 88.8% and 89.1% of samples, respectively, and A581G in 5.0% (**Table S1**). The *dhfr* triple-mutant IRN haplotype was observed in 28.4% of samples, and 86.4% of S108N carriers also harbored the K540E mutation (**Figure S6B**).

### Co-occurrence of 622I and mutations associated with reduced susceptibility to lumefantrine

We also assessed mutations in the context of partner drug sensitivity as continued efficacy of the partner drug is central to continued ACT efficacy. Mutations in the *mdr1* gene, including N86 (wild), 184F (mutant), and D1246 (wild) have been associated with decreased sensitivity to lumefantrine (commonly used partner drug in Ethiopia in ACT). The MDR1 NFD haplotype, which is linked to decreased lumefantrine susceptibility in vitro (Nzila et al. 2012), and the *mdr1* Y184F mutation alone were present in 98.2% (95% CI; 96.7-99.8%) of monoclonal parasites with a R622I mutation (**Figure 3A**). Furthermore, 78.2% (95% CI:73.5-83.0%) and 78.6% (95% CI:73.8-83.4%) of the R622I monoclonal parasites possess both the K76T and N75E mutations, as well as the N326S *crt* mutation, respectively (**Figure 3B**). Importantly, the prevalence of *crt* K76T mutation was more prevalent in parasites with the R622I mutation than in wild types (78.2% vs 67.6%, Pearson χ²=9.3273, p=0.002), indicating that R622I parasites select mutant genotype of *crt*. The co-occurrence of *k13* with *mdr1* and *crt* mutations in *P. falciparum* infections warrants close monitoring of efficacy of ACT and partner drugs in Ethiopia.

**Figure 3:**
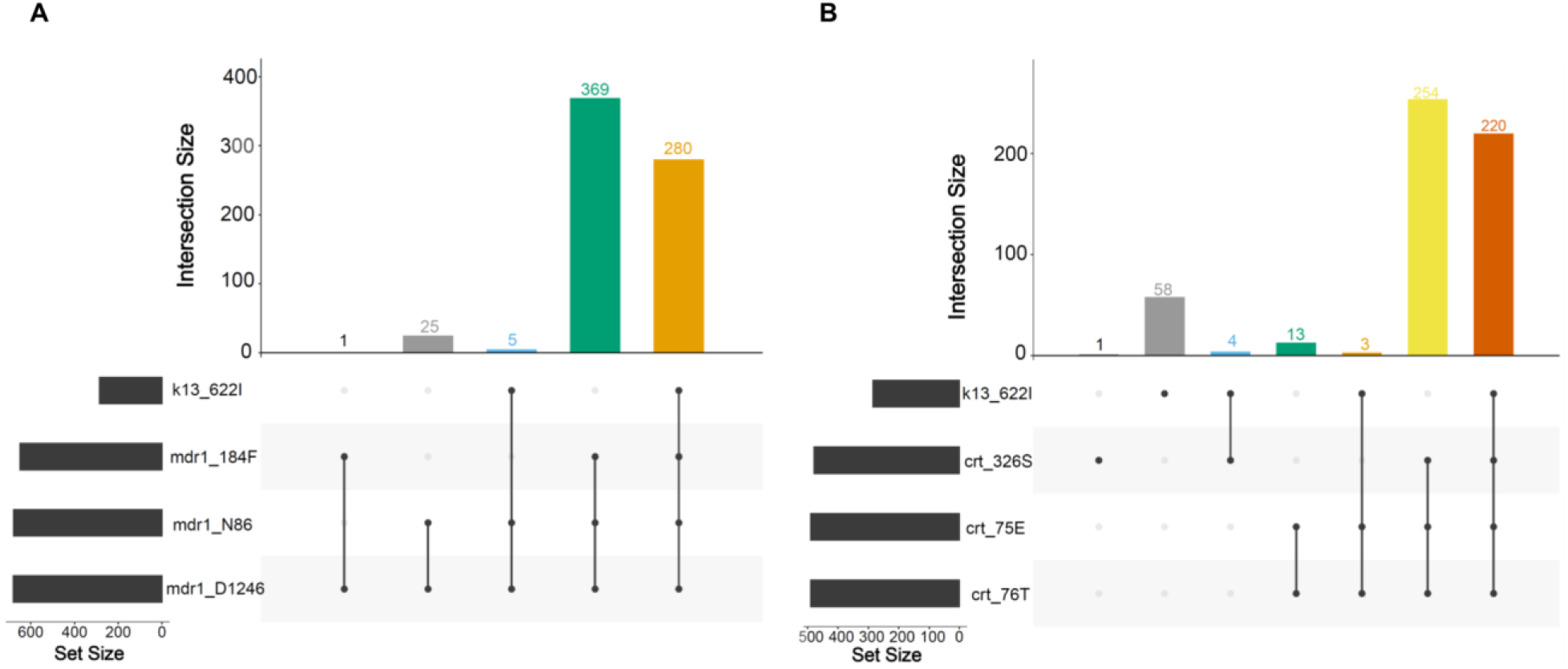
Prevalence of the *k13* R622I mutation and other partner drug-resistance mutations in monoclonal *P. falciparum* infections. **A**) Combinations of *k13* R622I and *mdr1* drug resistance mutations implicated in lumefantrine susceptibility, including the mdr1 NFD haplotype (N86 wild type, 184F mutant, and D1246 wild type). **B**) Combinations of *k13* R622I and *crt* drug resistance mutations implicated in aminoquinoline susceptibility, including *crt* (326S, N75E, and K76T). Only monoclonal samples (n=680) with genotypes for all loci were included.

### Genetic relatedness and connectivity of *k13* mutant parasites

To assess connectivity of the *k13* R622I mutant population at district and village levels, we analyzed pairwise relatedness using identity by descent (IBD) among monoclonal infections (88% of cases; **Figure S7**). The overall mean IBD was 0.264. In Gondar Zuria, R622I mutant pairs showed significantly higher IBD (0.369) than wild-type pairs (0.234; Student’s t-test p=3.29e-8). Similarly, in Tach Armachiho, IBD was higher among mutants (0.249) than wild types (0.172; Student’s t-test p=3.55e-3) (**Figure 4A**). Among R622I parasites, RDT-negative infections had greater relatedness (mean IBD=0.365) than RDT-positive ones (0.284; Student’s t-test *p*<0.001) (**Figure S8**). Highly related pairs (IBD≥0.95) showed clear clustering of R622I mutants by RDT status (**Figure 4B**).

**Figure 4.**
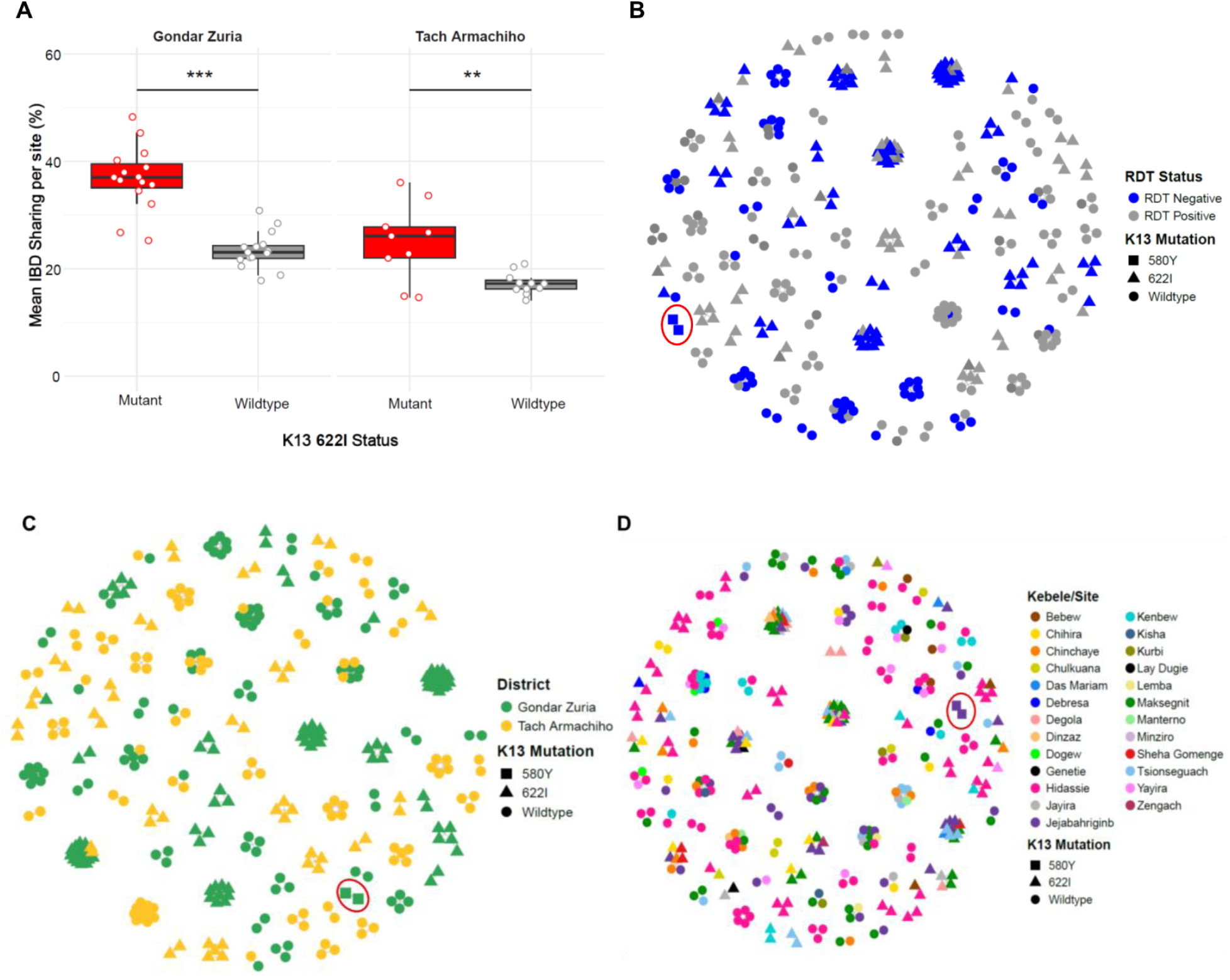
*k13* mutant and wildtype parasite relatedness. **A**) Pairwise IBD sharing between *k13* R622I mutant and wild-type *P. falciparum* parasites within kebeles across different study districts. Asterisks (*) denote level of statistically significant differences based on Student’s t-test: p < 0.01 (**), and p < 0.001 (***). Boxes indicate the interquartile range, the line indicates the median, the whiskers show the 95% confidence intervals and dots show within-kebele IBD values within each district. **B**) Network analysis of highly related parasite pairs (IBD≥0.95) by RDT test results shaped by *k13* mutation status. **C**) Network analysis of highly related parasite pairs (IBD≥0.95) by district shaped by *k13* mutation status. **D**) Network analysis of highly related parasites (IBD≥0.95) at Kebele (village) level shaped by *k13* mutation status. Each node identifies a unique isolate, and an edge is drawn between two isolates if they meet the IBD threshold. Within panels **B**, **C**, and **D**; the *k13* C580Y parasites, shaped as rectangles and encircled by red circles are clustered by RDT results, study district and village level, indicating their clonality. Color codes correspond to RDT results, districts and Kebeles (villages).

At a broader scale, *k13* R622I mutants exhibited clear clonal clustering within districts (**Figure 4C**). At the local level, parasites showed strong clustering within individual Kebeles, with some evidence of genetic connectivity between villages within the same district (**Figure 4D**). The two *k13* C580Y mutant parasites similarly showed tight clustering by RDT status, district, and village (**Figures 4B–D**, outlined in red circle).

### Monthly prevalence and clonality of *k13* mutations in northwest Ethiopia

We next examined the distribution of *k13* R622I over the course of the study, from November 1, 2022, to October 31, 2023. Although the differences over the months were not statistically significant (**Figure S9A and B**, Mann-Kendall trend, p>0.05), the peak *k13* R622I prevalence was 60.5% (95% CI 45.0-76.1%) in March. This peak follows the driest months of the year, with rainfall decreasing from November to February and resuming after March. The laboratory records from the health centers indicated that the number of *P. falciparum* cases was lowest in March. This pattern is also evident in the Tach Armachiho district (62.5%, 95% CI 38.8-86.2). In the Gondar Zuria district, the prevalence of the *k13* R622I mutation was slightly higher in September (67.6%, 95% CI 51.9-83.4), followed by March (59.1%, 95% CI 38.5-79.6). The lowest prevalence was observed in May for both the overall dataset (25.9%, 95% CI 16.2–35.8) and the Gondar Zuria district (27.1%, 95% CI 15.8–38.5), whereas in the Tach Armachiho district, the lowest prevalence occurred in April (18.2%, 95% CI 4.6–40.9). Throughout the year, the monthly prevalence of the R622I mutation remained consistently higher in the Gondar Zuria district than in Tach Armachiho, except in March **(Figure 5A**, **Table S4–S6**). Overall, this data suggests that *k13* R622I prevalence may be slowing and is not at a significant disadvantage in periods of low transmission.

**Figure 5:**
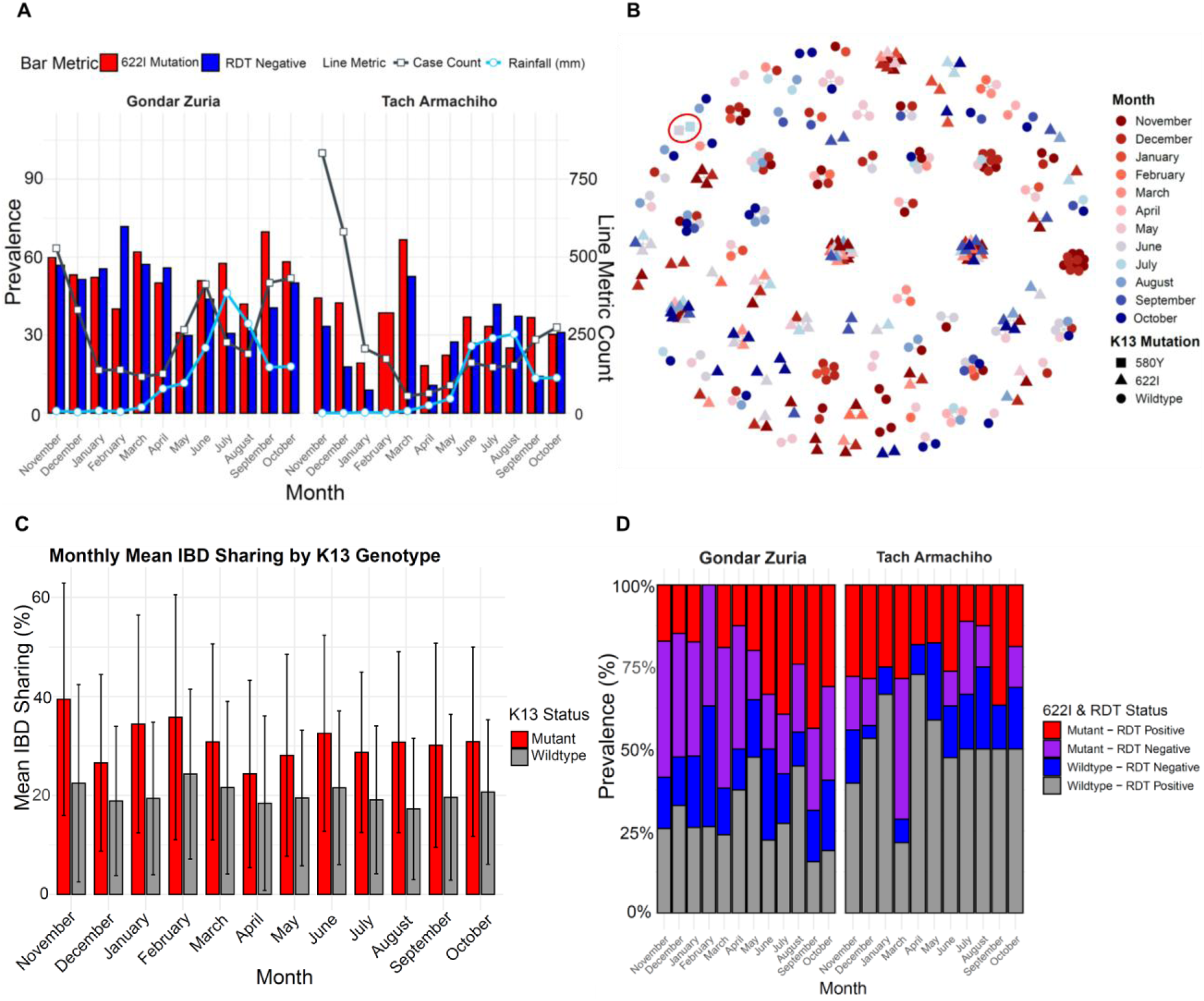
Monthly prevalence of the *k13* 622I mutation, RDT negativity, *P. falciparum* positivity and genetic relatedness in *P. falciparum* parasites. **A)** Monthly prevalence of *k13* 622I mutation and the rate of RDT negativity among *P. falciparum* isolates, along with rainfall and *P. falciparum* case count patterns across months per district. **B)** Network analysis of highly related monoclonal parasite pairs (IBD≥0.95) by month, where each node represents a unique isolate, and edges are drawn between isolates meeting the IBD threshold; isolates without such connections are not shown, and color codes correspond to different months. Within this panel, C580Y parasites, which are depicted as rectangles and encircled in red, share IBD≥0.95 indicating their clonality and persistence between consecutive months. **C)** Monthly mean pairwise IBD of *P. falciparum* isolates by *k13* genotype status. Error bars represent the 95% confidence intervals. **D)** Distribution of 622I mutation by RDT result status across months of the study districts.

IBD network analysis showed that the R622I mutants are closely related and maintain population connectivity throughout the study period, whereas the wildtype samples exhibit greater differentiation over time. This is demonstrated by the fact that the R622I mutants from different months form larger clusters, unlike the wildtype samples which form many small clusters from adjacent months (**Figure 5B**) consistent with a lower mean pairwise IBD compared to the mutants across all months (**Figure 5C**). RDT-negative and R622I mutant parasites were detected throughout all months in the Godar Zuria district and in most months in the Tach Armachiho district. It’s also interesting to note that March was the month when the co-occurrence of RDT-negativity and the R622I mutation peaked in both districts (**Figure 5D**), suggesting RDT negativity may aid in avoiding treatment during low transmission periods.

## Discussion

In-depth sampling and analysis of two districts over a year in the region where *k13* R622I mutation was first identified in Africa (Bayih et al., 2016) confirm rising prevalence and emergence of other WHO-validated and candidate ART-R mutations, including C580Y. The 622I mutation was detected overall in nearly half of all infections, with the majority (51.4%) in the Gondar Zuria, a highland district with lower transmission intensity. It was distributed across virtually all surveyed Kebeles and showed higher genetic similarity among mutant parasites, pointing to recent clonal expansion and rapid spread. However, there was no significant increase in prevalence over the year suggesting the rate of increase may be slowing or plateauing. R622I was positively associated with HRP2-based RDT negativity, raising serious concerns about missed diagnoses and untreated infections in the past, prior to Ethiopia’s recent change to non-HRP2-based RDTs. The greater relatedness and association with RDT negativity suggest there could be a propensity now for clonal transmission of *k13* R622I, *hrp2* deletion, and *hrp3* deletion which occurs on 3 separate chromosomes. Additionally, we found that R622I mutation persisted throughout the year and may even potentially increase during the dry season, when case numbers were lowest. This temporal pattern, coupled with high genetic relatedness among R622I mutants across months, suggests that these parasite populations can persist and expand across seasons.

In relation to past reports, we find that *k13* R622I prevalence has rapidly increased in northern Ethiopia, at 51.4%, and 34.5% in the two districts deeply sampled. This marks a striking increase from earlier reports: 2.4% in 2014 (Bayih et al., 2016), 9.5% in 2017 (Alemayehu et al. 2025), 10% in 2018 (with site-level prevalence at Tegede reaching 28.3%) (Fola et al., 2023) and 19.44% in 2021 (Kamaliddin et al. 2024) in the Amhara region. These trends suggest mounting selective pressure, most likely driven by widespread ACT use. A similar pattern has been observed in Eritrea, where the R622I mutation was detected in 13.3% of samples, with prevalence increasing from 8.6% in 2016 to 21.0% in 2019, reaching 29.1% in one site and over 20% in multiple other districts (Mihreteab et al. 2023). Notably, in the same study R622I was more common in patients with persistent Day 3 parasitemia, though *in vitro* assays showed it conferred only low-level artemisinin resistance compared to the C580Y mutation. Such functional validation has not yet been performed for R622I genotypes circulating in Ethiopia, which may harbor distinct genetic backgrounds.

The consistently high prevalence across nearly all sampled sites, particularly in lower transmission settings, indicates that artemisinin-resistant parasites are now firmly established in northern Ethiopia. The higher prevalence of mutant parasites in the lower transmission district aligns with expectations: reduced within-host competition from wild-type parasites (Bushman et al. 2018) and a greater likelihood of treatment exposure (Mihreteab et al., 2023). Supporting this, R622I mutants exhibited significantly higher mean pairwise IBD values than wild-types, consistent with increased selfing and clonal propagation under reduced transmission (Anderson et al., 2000; Camponovo, Buckee, and Taylor, 2023). These patterns mirror previous findings of clonal R622I transmission in Ethiopia (Fola et al., 2023; Brhane et al., 2024). However, the detection of a few highly related parasite genotypes shared across districts suggests occasional importation or human-mediated movement (Haile, Lemma, and Weldu, 2017). Together, these findings indicate that R622I is expanding through both local clonal amplification and limited regional connectivity.

In addition to the widespread *k13* R622I mutation, we also detected C580Y mutation, a WHO-validated marker of high-level artemisinin resistance and the most globally recognized variant associated with treatment failure in SEA (Ariey et al. 2014; Ashley et al. 2014). While C580Y has been previously reported outside SEA including in Guyana (Chenet et al. 2016), Papua New Guinea (Miotto et al. 2020), Ghana (Aninagyei et al. 2020), and Nigeria (Martín Ramírez et al. 2025) this represents the first confirmed detection of C580Y in the Horn of Africa. Unlike the unvalidated Ghana and Nigeria reports, our findings are supported by MIP and WGS sequencing and show that the two C580Y-positive samples from Gondar Zuria district, the same Kebele were monogenomic and clonal (IBD=1), suggesting a recent and possibly local emergence. Phylogenetic and flanking haplotype analysis based on whole-genome ONT sequencing revealed no significant relatedness to SEA C580Y lineages, indicating de novo emergence within Africa. Notably, one of the patients had been treated with artemether-lumefantrine two weeks prior to sample collection, suggesting potential treatment failure. The clonal nature of these infections, their geographic clustering, and signs of local transmission echo the early expansion of the locally emerged *k13* R561H variant in Rwanda (Uwimana et al. 2020). These findings highlight the urgent need for strengthened genomic surveillance and targeted ART-R containment efforts in East Africa to monitor and mitigate the spread of high-level artemisinin resistance (Neafsey and Volkman 2017; Auburn and Barry 2017).

Another key new observation from this study is the emerging association between the *k13* R622I mutation and HRP2-based RDT negativity, marking a shift from earlier reports where a relationship between *k13* 622I and hrp2/3 deletion was not observed in Ethiopia (Fola et al., 2023). Parasites carrying the R622I mutation were significantly more likely to test negative by HRP2-based RDTs, with 48.3% of R622I infections being RDT-negative compared to only 30.7% of wild-type infections (p<0.05). This pattern of strong clustering by RDT status suggests co-occurrence of R622I mutations and *hrp2/3* deletions consistent with recent findings of high co-occurrence of R622I and *hrp2/3* deletions in Eritrea (Mihreteab et al., 2023). Although we did not directly confirm deletions with molecular assays in this study, the lack of significant differences in parasite densities between RDT-positive and RDT-negative samples suggests that diagnostic failure is likely due to genetic deletions rather than low parasitemia (Feleke et al., 2021). In areas like the Horn of Africa, where test-and-treat strategies are widely implemented, this convergence likely reflects intense selective pressure that favors parasites able to escape both diagnosis and clearance (Fola et al., 2023; Mihreteab et al., 2023). Encouragingly, the recent shift in Ethiopia and Eritrea toward non-HRP2-based RDTs may help mitigate the selective advantage of HRP2-deleted parasites, potentially slowing the future spread of dual-resistant strains.

Temporal trends observed in this study, based on continuous year-round sample collection, provide important insights into the epidemiological dynamics of artemisinin-resistant *P. falciparum* in northern Ethiopia. While the overall prevalence of the *k13* 622I mutation remains alarmingly high, its rate of increase appears to be slowing, suggesting a potential plateau in its expansion. This decline may reflect a shift in the selective landscape, possibly due to saturation of the mutation in the parasite population or localized changes in drug pressure caused by ongoing conflict that disrupted malaria control efforts (Mertens 2024; Yu et al. 2024). However, our ability to assess long-term trends is limited by the fact that data were collected over only a single year. Notably, we found compelling evidence that R622I mutant parasites persist through the dry season, a period typically associated with reduced transmission, indicating their ability to maintain infections despite lower vector activity. Such persistence suggests either longer infection durations, low-density chronic carriage, or ongoing low-level transmission despite ecological constraints which needs further investigation including asymptomatic malaria cases as our current study only focuses on symptomatic cases. Moreover, prevalence of mutation increased immediately after the rainy season, when transmission began, and drug use increased as seen in other studies (Ord et al. 2007; Asih et al. 2009). These findings underscore that the R622I mutation is not only entrenched but may also be adapting to seasonal fluctuations in transmission intensity, thereby enhancing its potential for long-term establishment and year-round transmission.

Despite the increasing prevalence of *k13* mutations, artemisinin-based combination therapy (ACT) continues to demonstrate apparent clinical effectiveness in Ethiopia (Daka et al. 2024; Deressa et al. 2017), likely due to the potent activity of partner drugs used alongside artemisinin derivatives (WHO, 2018). However, ART-R can prolong parasite survival following treatment, increasing the duration during which parasites are exposed solely to the partner drug, thereby elevating the risk of selecting for partner drug resistance. ART-R has been shown to accelerate the spread of such resistance (Imwong et al., 2017). In our study, we observed extremely high frequencies of *mdr1* mutations associated with reduced lumefantrine sensitivity, with the NFD haplotype (N86 wild-type, 184F mutant, D1246 wild-type) present in 98.2% of 622I mutant monoclonal infections and reaching near-fixation levels overall (96.8%–100%). This raises significant concern for declining lumefantrine efficacy (Grossman et al. 2023; Halsey and Plucinski 2023), and suggesting strong regional selection pressure driven by widespread ACT use (Okell et al., 2018). The co-occurrence of high-frequency partner drug resistance markers and predominantly monogenomic infections highlights a growing risk of reduced ACT effectiveness in Ethiopia, a concern echoed by recent reports of declining treatment efficacy in parts of Africa (Arya et al., 2021, van Schalkwyk et al. 2024).

While our deep sampling across two ecologically distinct regions offers a valuable snapshot of resistance patterns, several limitations remain. First, the lack of clinical outcome data and absence of in vitro or in vivo efficacy assessments limit our ability to directly associate the observed molecular markers with treatment failure. Second, site-level sample sizes were relatively low during the dry season, when malaria transmission is minimal, which may have reduced our power to detect spatial and seasonal variation. Finally, as the study covers only a single year, we were unable to capture longitudinal trends or assess year-to-year changes in resistance dynamics.

In conclusion, our study highlights a growing threat to the efficacy of artemisinin-based combination therapies (ACTs) in northern Ethiopia. We report a sharp rise in the prevalence of the *k13* R622I mutation, marked by clonal expansion and sustained year-round transmission. Notably, we document the first confirmed detection of the *k13* C580Y mutation in Ethiopia/Horn of Africa, emerging independently of SEA lineages. Resistance appears compounded by the frequent co-occurrence of artemisinin-resistant alleles with the MDR1 NFD haplotype, which has been associated with reduced lumefantrine sensitivity. Furthermore, the significantly higher rate of HRP2-based RDT negativity among 622I-carrying parasites reveals a troubling convergence of drug and diagnostic resistance, posing critical challenges for malaria control and elimination efforts in the region.

## Methods

### Study site

The study was conducted from November 01, 2022 to October 31, 2023 in Gondar Zuria and Tach Armachiho districts, Amhara region, northwest Ethiopia (**Figure 1A**). The Gondar Zuria district is about 45 km away from Gondar town, which is 686 km far from the capital city (Addis Ababa), Ethiopia. The area of Gondar Zuria District is 1108.53 km^2^, with an estimated population size in 2023 of 252,091. It has more than 35 smallest administrative units called Kebeles and is bordered on the south by the Debub Gondar Zone, on the southwest by Lake Tana, on the west by Dembiya, on the north by Lay Armachiho, on the northeast by Wegera, and on the southeast by Mirab Belessa. Its climate is Woina Dega (subtropical zone) with an altitude ranging between 1750 and 2600 meters above sea level. It has an average annual temperature of about 25.1 degree Celsius with annual rainfall between 719.9 and 1831.9 millimetres. During the study period, the district had 8 health centers and 1 hospital. The Tach Armachiho district is located approximately 65 kilometers from Gondar town and 796 kilometers from Addis Ababa. The district is divided into 24 Kebeles and is bordered by Tegedie to the north, Lay Armachiho and Chilga to the south, Metema to the southwest, Mirab Armachiho to the west, and Dabat districts to the east. Its total area is 2,710.41 km^2^ and according to the 2023 estimation, the district’s total population was 121,597. It is a low land area (tropical zone) with an altitude of below 1500 meters above sea level. The mean minimum and maximum temperatures recorded annually are 30°C and 42°C, respectively. The district has a unimodal rainfall pattern and extends from June to the end of September months (unpublished district Agriculture office document, 2019). Throughout the study period, the district was equipped with 6 health centers and 1 hospital. Over the last of two to three years, there has been a rise in the number of malaria cases reported by the health department of the respective districts. The Ethiopian Federal Ministry of Health’s categorization of malaria transmission intensity places Tach Armachiho district in the high transmission zone, while Gondar Zuria district is identified as having a moderate transmission level.

### Sample collection and testing malaria cases

Health facility based cross sectional study was conducted over a year. The sources of population were patients who were sent to the laboratory for malaria screening in the selected health centers from the study districts. Maksegnit and Sanja health centers, serving as central hubs for all villages, were chosen from Gondar Zuria and Tach Armachiho districts, respectively. Microscopically confirmed *P. falciparum* infected patients, ≥ 2 years of age & signed informed written consent were included in the study as study population. Patients who were not volunteers, have bleeding disorder and with severe malaria symptoms were excluded from the study. All *P. falciparum* positive patients (n=903, GZ=500, TA=403) who fulfilled the inclusion criteria within the study period were included in the study. Finger prick blood samples (∼375 µL) were collected using EDTA-coated microtainer tubes.

Thin, and thick smears were made for confirmation and quantification of parasite densities by expert microscopists. Both thick and thin blood films were prepared from finger prick blood on a single slide and stained using Giemsa solution as described before. Parasitic stage densities were determined using previously used protocol (Herman et al. 2019). The degree of parasitemia was categorized as low, moderate, and high parasitemia when the count was between 1-999, 1000-9999, and ≥10,000 parasites/μl, respectively. Rapid diagnostic test (RDT) targeting *P. falciparum* histidine-rich protein 2 (HRP2) was also done. RDTs are chromatographic tests for in vitro diagnosis of malaria parasites. They contain a membrane strip pre-coated with a monoclonal antibody against malaria parasite antigens as a single line across the test strip conjugated to a signal, typically colloidal gold (Maltha, Gillet, and Jacobs 2013). The monoclonal antibodies were species specific. The RDTs were performed according to the manufacturer’s instructions. Data for all patients screened for malaria from November 1, 2022, to October 31, 2023, at the two health centers were extracted from laboratory log books. Rainfall data were obtained from the CHIRPS: Rainfall Estimates from Rain Gauge and Satellite Observations v3.0 dataset (Funk et al. 2015) using the longitude and latitude coordinates of each sampled kebele. The average rainfall values for each sampled kebele from each district were used to represent the rainfall pattern of the study districts.

### DNA extraction and MIP sequencing

Dried blood spots (DBS) were prepared on filter papers (Whatman, Maidstone, UK) for molecular assays as previously described (Brhane et al. 2024). In brief, the DBS samples were air dried and kept in zip-locked plastic bags containing self-indicating silica gel desiccant beads (Sigma-Aldrich) and transported to University of Gondar and stored at −20°C freezer until they were transported to Brown University, Rhode Island, USA. DNA was extracted from DBS (one full 6mm DBS spot, typically involving 2 or 3 punches) using Chelex-Tween as previously described (Teyssier et al. 2021).

Five microliters of extracted DNA were used for each of the MIP captures using two panels: one covering key *P. falciparum* drug resistance genes and mutations (DR23KE), including *k13*, *mdr1, crt, dhfr,* and *dhps* genes, and a newly designed panel of 2128 MIPs (IBC2FULL) targeting common SNPs (>5%) across *P. falciparum* genome (Niaré et al. 2025). MIP capture and library preparation were performed as previously described (Verity et al. 2019; Fola et al. 2023). Sequencing was conducted using an Illumina NextSeq 550 instrument (150 bp paired-end reads) at Brown University (RI, USA). For samples with newly detected *k13* C580Y mutation in Ethiopia, an additional MIP capture was done using the same MIP panel and resequenced to a high depth to confirm the mutations. Controls for each MIP capture and sequencing included genomic DNA from serial dilution of lab strain 3D7 as well as no template and no probe controls.

### MIP data analysis

Processing of sequencing data and variant calling was done using MIPtools (v0.19.12.13; https://github.com/bailey-lab/MIPTools), a suite of computational tools designed to process sequencing data from MIPs. Raw reads from each MIP, identifiable using unique molecular identifiers (UMIs), were used to reconstruct sequences using MIP Wrangler, and variant calling was performed on these samples using Freebayes (Garrison and Marth 2012). Variants were annotated using the 3D7 v3 reference genome. For the genome-wide MIP panel, variants were quality filtered by removing those with less than 3 UMIs within a sample and less than 10 UMIs across the entire population. To reduce false positives due to PCR and alignment errors, the alternative allele (SNP) must have been supported by more than one UMI within a sample, and the allele must have been represented by at least 10 UMIs across the entire population. Biallelic, variant SNP positions were retained for downstream analyses. Moreover, SNPs with more than 50% missing data, followed by samples with less than 50% data, were removed from all downstream analyses.

### Estimation of drug resistance prevalence

The panel for detecting drug resistance included key and known SNP in genes such as *k13, crt, dhfr, dhps, mdr1*, along with other candidate drug resistance genes, as previously mentioned (Aydemir et al. 2018). The rate of mutations associated with drug resistance was determined by dividing the number of infections carrying these mutations by the total number of successfully sequenced infections, then multiplying by 100 to get the prevalence. This filtering process of variants and estimation of drug resistance mutations utilized the miplicorn R package version 0.2.90 (https://github.com/bailey-lab/miplicorn), and the vcfR R package version 1.13.0 from (https://github.com/daniellyer/vcfR). Graphs displaying combinations of drug-resistant mutations were generated and displayed using the ‘UpSetR’ Package in R version 1.4.0 from R Project Repository (Conway, Lex, and Gehlenborg 2017). Testing for district-level temporal trends in 622I prevalence and RDT-negativity rate was performed using the “Kendall” package in R version 2.2.1 from the R Project Repository.

### Complexity of infection and parasite relatedness analysis

Using the final variant set (n=2472 SNPs) spread across the genome, we calculated the complexity of infection (COI) for each sample utilizing THE REAL McCOIL categorical method (Chang et al. 2017). This method converts heterozygous SNP data into reliable estimates of allele frequencies, achieved through Markov chain Monte Carlo (MCMC) methods. We employed the hmmIBD method to estimate the genetic relatedness among parasite strains samples (Schaffner et al. 2018). Samples were considered to be identical-by-descent if they shared IBD≥0.95 based on a minimum of 50 informative SNPs. We used likelihood-ratio test statistics to test the null hypothesis that two samples are unrelated (*H0*: IBD = 0) at significance level *α* = 0.05 (with the procedure adjusted for a one-sided test). We assessed mean IBD and the number of related samples for different groups and performed permutation tests to determine which combinations had higher mean IBD and more related samples than expected by chance. Relatedness networks of parasites were created using the R *igraph* package.

### Long-read whole-genome sequencing and haplotype analysis

Whole-genome sequencing using Oxford Nanopore Technology (ONT) P2 Solo was done on 2 DNA samples (samples with *k13* C580Y mutations based MIP sequencing) to confirm the presence of the mutation within kelch13 propeller domain region of *k13* gene and to assess whether the mutation is locally emerged or imported using extended haplotypes analysis. To get enough template for nanopore sequencing, samples were subjected to selective whole-genome amplification (sWGA) as described with minor modifications (Zuckerman & Fola, *in preparation*). In brief sWGA selectively amplifies the target genome (here *Plasmodium falciparum* DNA) over background DNA (human genome) using a pool of primers designed to amplify frequently occurring motifs of short nucleotides in *P. falciparum* reference genome. The sWGA experiment was performed in two steps. Combining: 8 ul DNA sample, 0.25 µL (20uM concentration of each primer in the pool), 0.5 µL of 10X ThermoFisher EquiPhi29 reaction buffer, and 1.25 µL nuclease-free water, the resulting 10uL was denatured for 3 minutes at 95°. 2^nd^ The denatured product was then mixed with 1 ul (10 units) of EquiPhi29 DNA polymerase, 2 µL reaction buffer 0.2 µL of 100µM MDT, 2µL of 10mM dNTPs, and nuclease-free water to make a total pool volume of 20µL and isothermally amplified 45°C for 3 hours, then 65°C for 10 minutes to suspend further enzyme activity. Amplification success was validated with measuring DNA quantity before and after enrichment using Qubit. The sWGA product was then prepared for sequencing on the Nanopore P2 Solo using the Native Barcoding Kit V14 (SQK-NBD114.24) and the Ligation Sequencing gDNA protocol.

For comparison of C580Y haplotypes, publically-available whole-genome sequencing from SEA with C580Y mutation (n=41) (MalariaGEN et al. 2023). High-accuracy duplex base-calling model was done by using Dorado to increase accuracy and alignment was done with minimap tools followed by clair3 variant calling (Zheng et al. 2022). The *k13* flanking haplotypes for C580Y Ethiopian and SEA strains were visualized by merging variant calls at shared sites and applying a custom plotting script. Although coverage was poor for one of the two long-read sequenced Ethiopian C580Y mutant parasites, the other sample had sufficient coverage and was used for phylogenetic analysis. This well-covered sample was confirmed to have a distinct flanking haplotype in the region surrounding the mutation of interest.

### Ethical clearance

Ethical clearance with reference number of VP/RTT/05/814/2022 was obtained from the institutional review board (IRB) of University of Gondar. Permissions from the respected Health Offices of the study districts and village administration were also obtained. Informed consent was obtained from all adult participants or from guardians in the case of children, while assent was also obtained from children aged 12 to 17 years prior to sample collection. When the study was being carried out, all appropriate safety measures and ethical methods were taken into account for both the participants and the researchers. Only authorized staff members have access to the protected site where all screening and case record forms are stored. The computer-based data entry employed unique numerical identifiers. Genotyping work in Brown University was regarded as non-human subject research. For reporting individual level genotypes for publishing, only de-identified samples and aggregated clinical data were utilized.

### Data and codes availability

All sequencing data from the current study are available under accession numbers SAMN48791106–SAMN48792008 at the Sequence Read Archive (http://www.ncbi.nlm.nih.gov/sra). The associated BioProject can be found under accession number PRJNA1269601 (https://www.ncbi.nlm.nih.gov/bioproject/). The data includes de-identified patient clinical information, a data dictionary, and drug resistance genotypes, all of which are available in the supplementary tables. All code and input files to generate the main figures for this manuscript are available on Github (https://github.com/gtollefson/Northern_Ethiopia_Pf_ARTR_2025_Project).

### Author contributions

AJZ, AAF, PK, AH, MA and JAB conceived the study. AJZ led patient recruitment and sample collection. AJZ, AL, RC. OT, BGB, AA and AAF performed laboratory work. AJZ, GT, JM, KN and AAF led genetic data analysis. AJZ and AAF wrote the first draft of the manuscript. JAB and JBP supported genetic data analysis and interpretations. All authors edited the manuscript and approved the final version before submission.

## Data Availability

All data produced are available online at

https://www.ncbi.nlm.nih.gov/sra

https://www.ncbi.nlm.nih.gov/bioproject/

https://github.com/gtollefson/Northern_Ethiopia_Pf_ARTR_2025_Project

## Acknowledgments

We are also grateful to all study participants and data collectors.

## Supplementary materials

### Supplementary Figures

**Figure S1.**
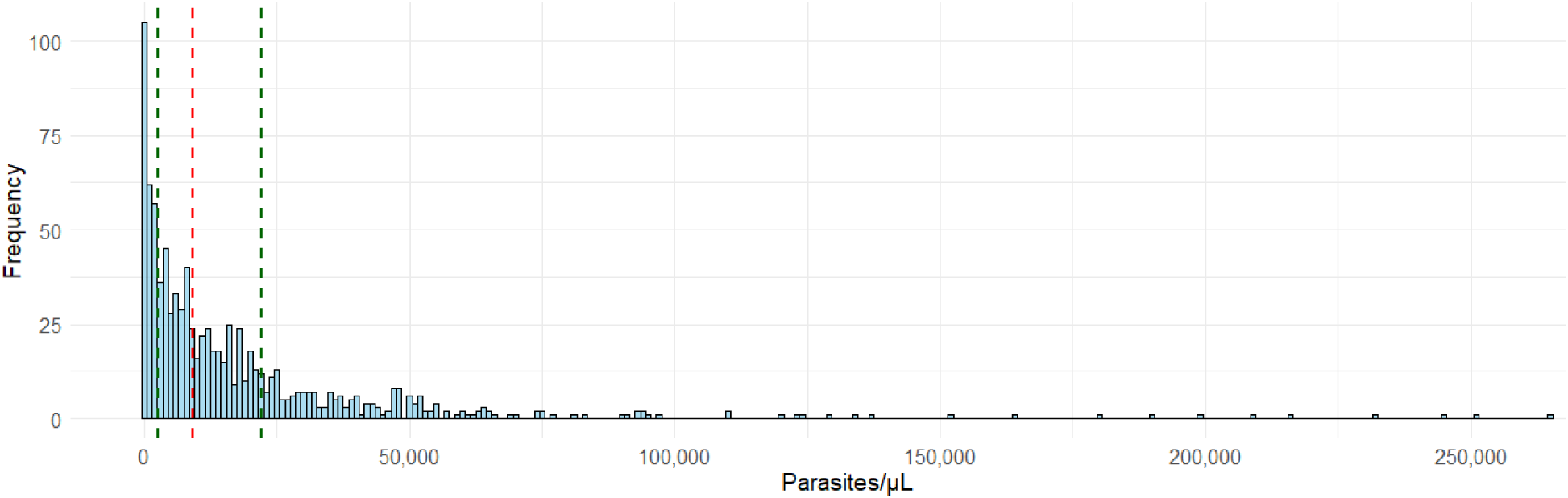
Histogram showing the distribution of *P. falciparum* parasite density (measured by microscopy) in samples from Northwest Ethiopia. The median density was 9,121 parasites/μL (red line), with the 25th and 75th percentiles at 2,519 and 22,160 parasites/μL, respectively (green lines). One outlier sample with 495,686 parasites/μL was excluded to enhance clarity of the distribution.

**Figure S2.**
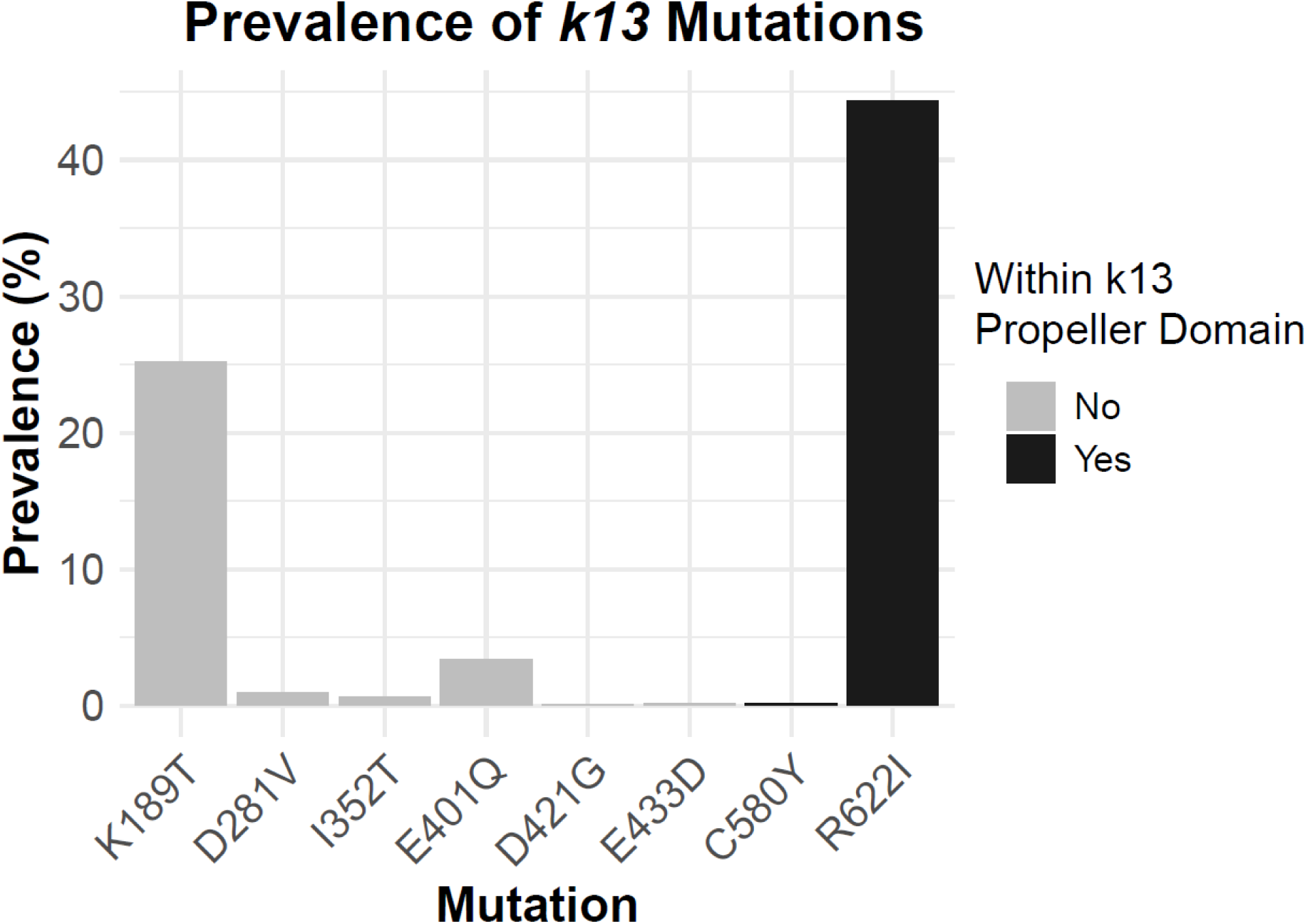
Prevalence of *k13* gene mutations in samples collected between November 1, 2022, and October 31, 2023, from Gondar Zuria and Tach Armachiho districts in Northwest Ethiopia.

**Figure S3.**
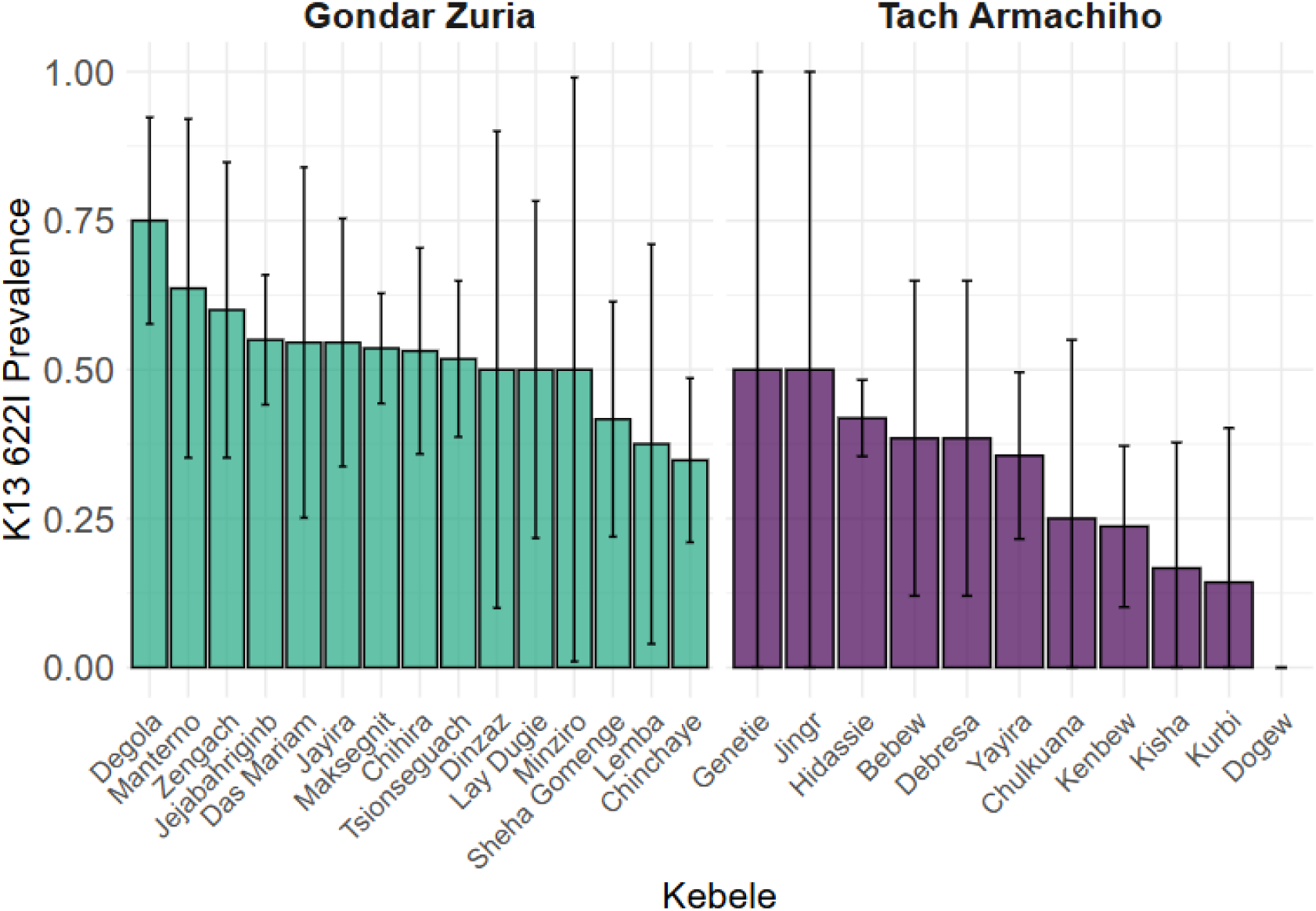
Prevalence of the *k13* R622I mutation across sampled kebeles (villages) within each study district. Error bars represent the 95% confidence intervals of the prevalence estimates.

**Figure S4.**
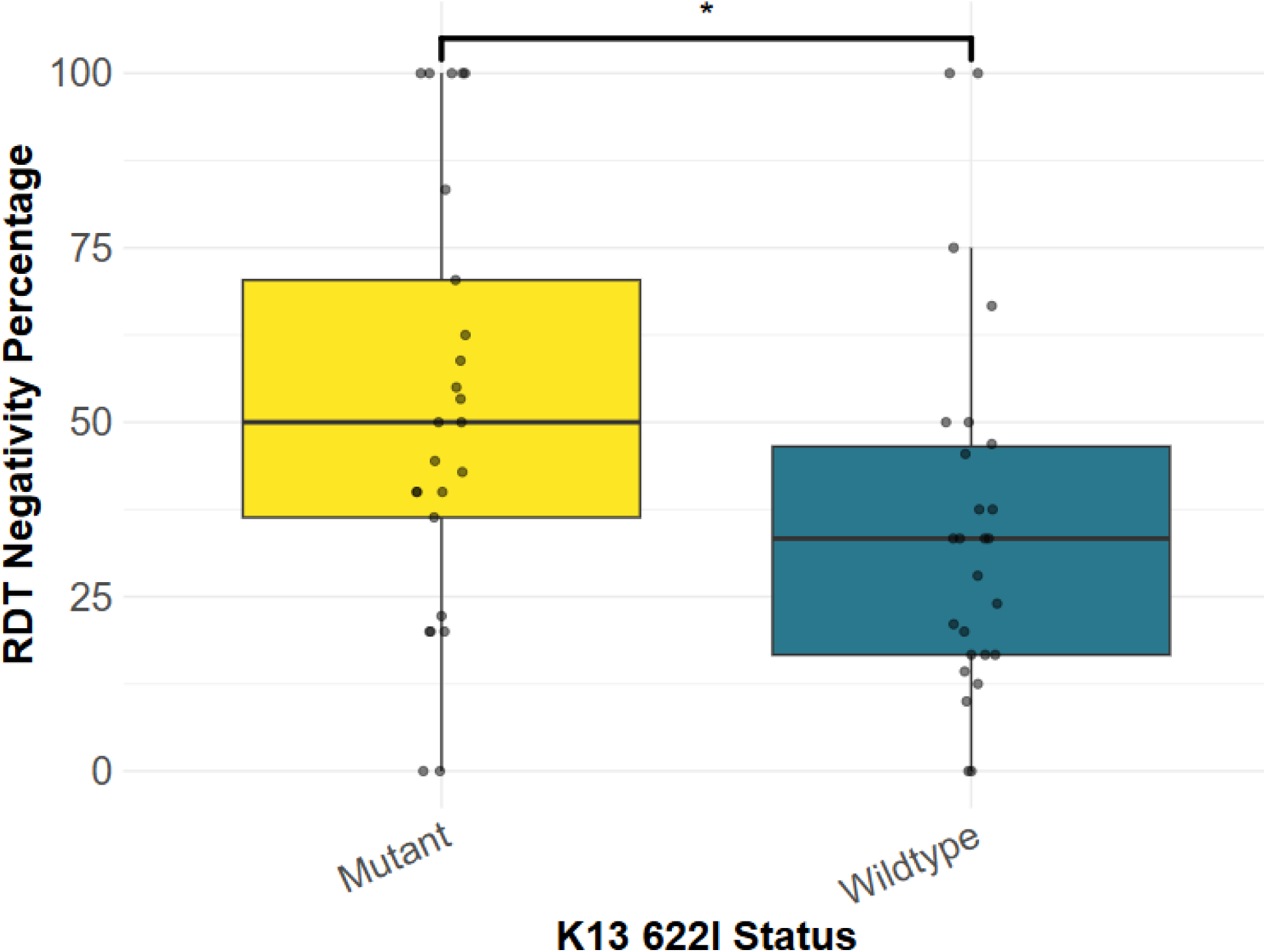
Prevalence of *pfhrp2*-based RDT negativity stratified by *k13* 622I mutation status. The boxplot displays the median (center line), with the box edges indicating the 25th and 75th percentiles. .Student’s *t*-test: *, *p* < 0.01.

**Figure S5.**
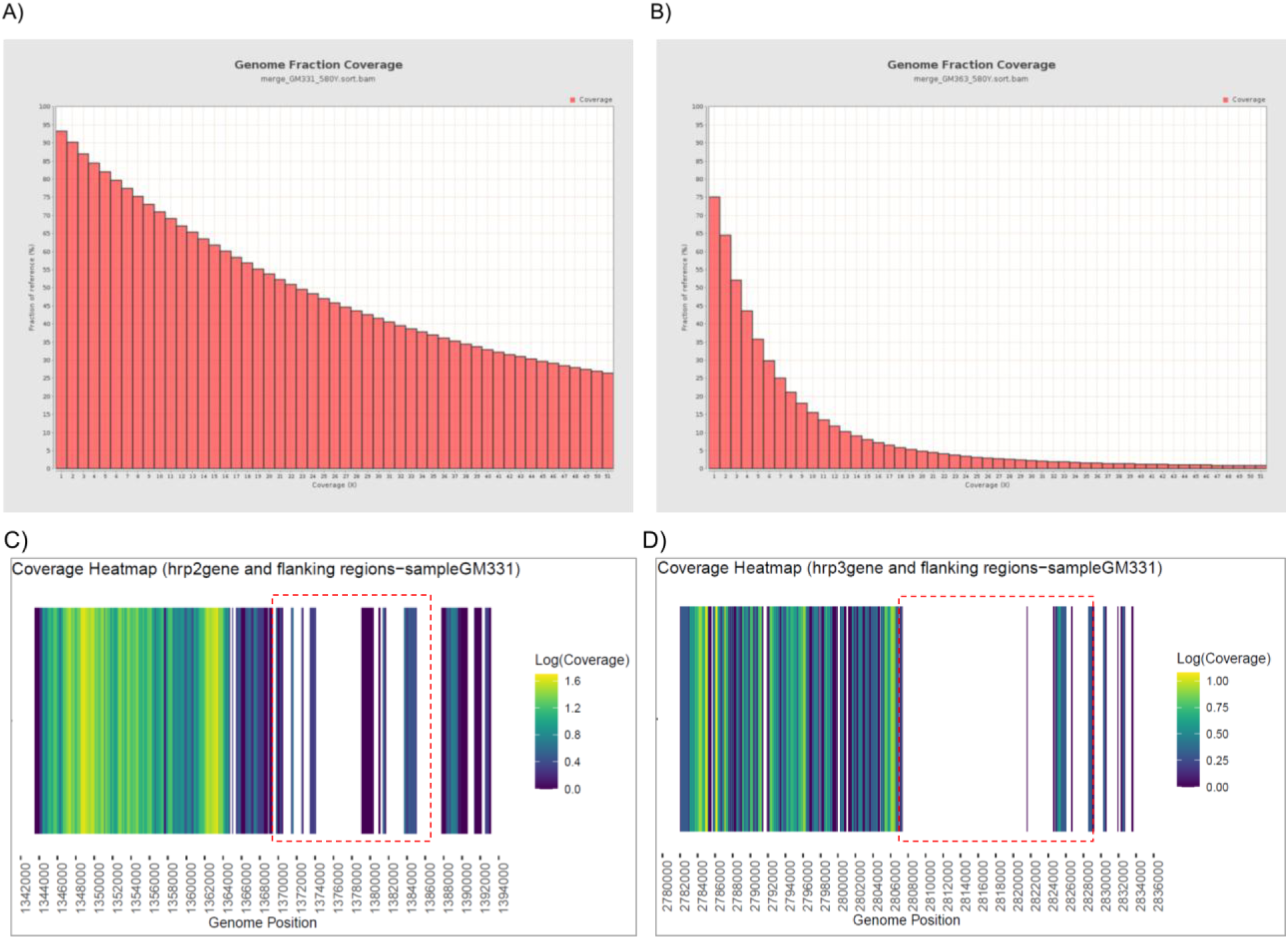
**Genome coverage and evidence of *hrp2/3* gene deletions in C580Y mutant parasites based on whole-genome sequencing.** (A) Genome coverage of C580Y mutant from sample GM331, with 82% of the genome covered at ≥5× depth. (B) Genome coverage of C580Y mutant from sample GM363, with 36% of the genome covered at ≥5× depth. (C) The BAM file coverage plot shows evidence of hrp2 gene deletion in the C580Y mutant, indicated by the broken red rectangular regions. (D) The BAM file coverage plot shows evidence of hrp3 gene deletion in the same parasite, indicated by the broken red rectangular regions. The white spaces C and D represent genomic regions with a complete absence of sequencing reads, consistent with gene deletion.

**Figure S6.**
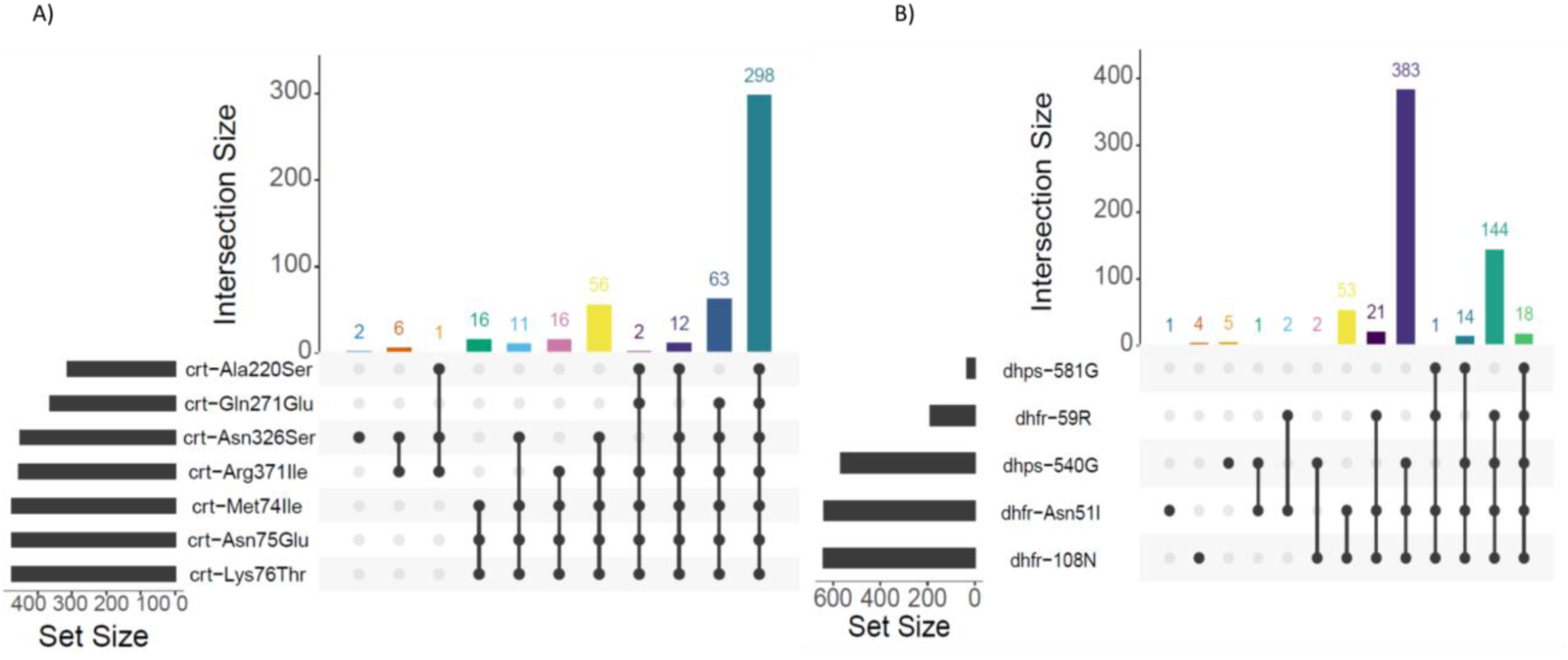
**Prevalence of mutations in key antimalarial drug resistance genes.** (A) Combinations of *crt* gene mutations associated with drug resistance. (B) Combinations of *dhps* and *dhfr* gene mutations linked to antifolate resistance.

**Figure S7.**
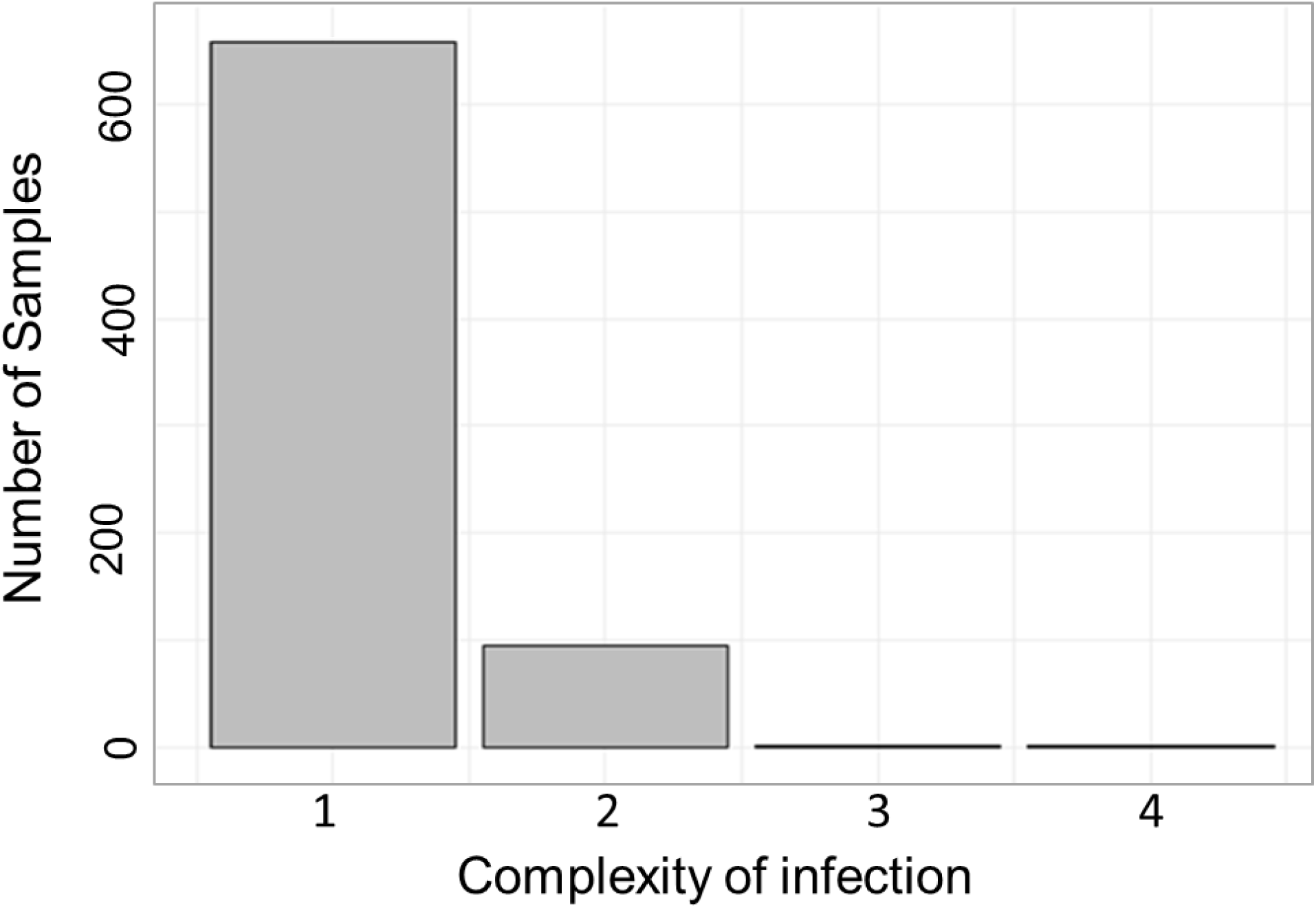
Frequency of samples by complexity of infection (COI) category, with nearly 88% classified as monoclonal infections.

**Figure S8.**
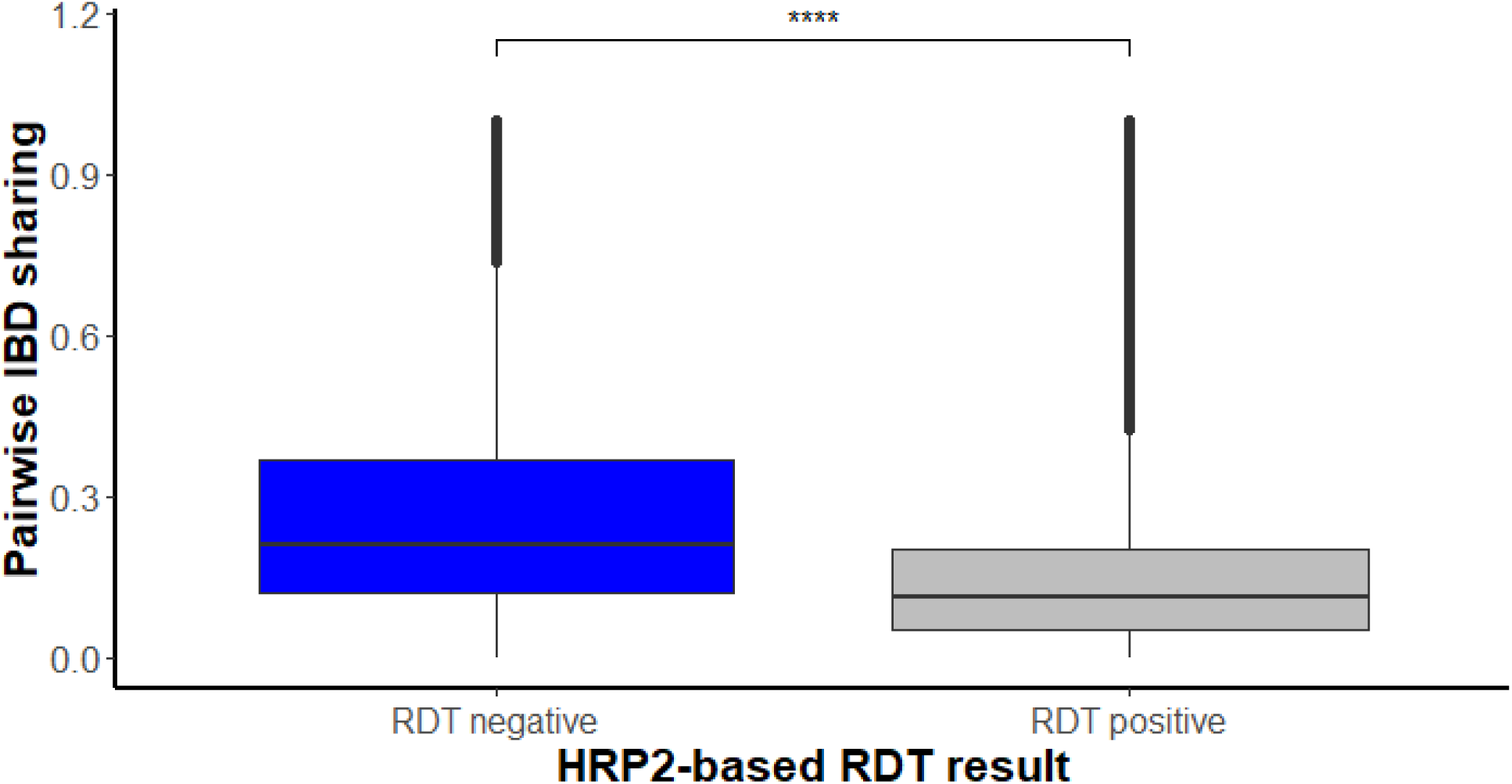
Mean identity-by-descent (IBD) sharing among *k13* 622I parasites, stratified by *pfhrp2*-based RDT result. IBD sharing was higher among RDT-negative samples (Student’s *t*-test; **** (*p* < 0.0001)

**Figure S9.**
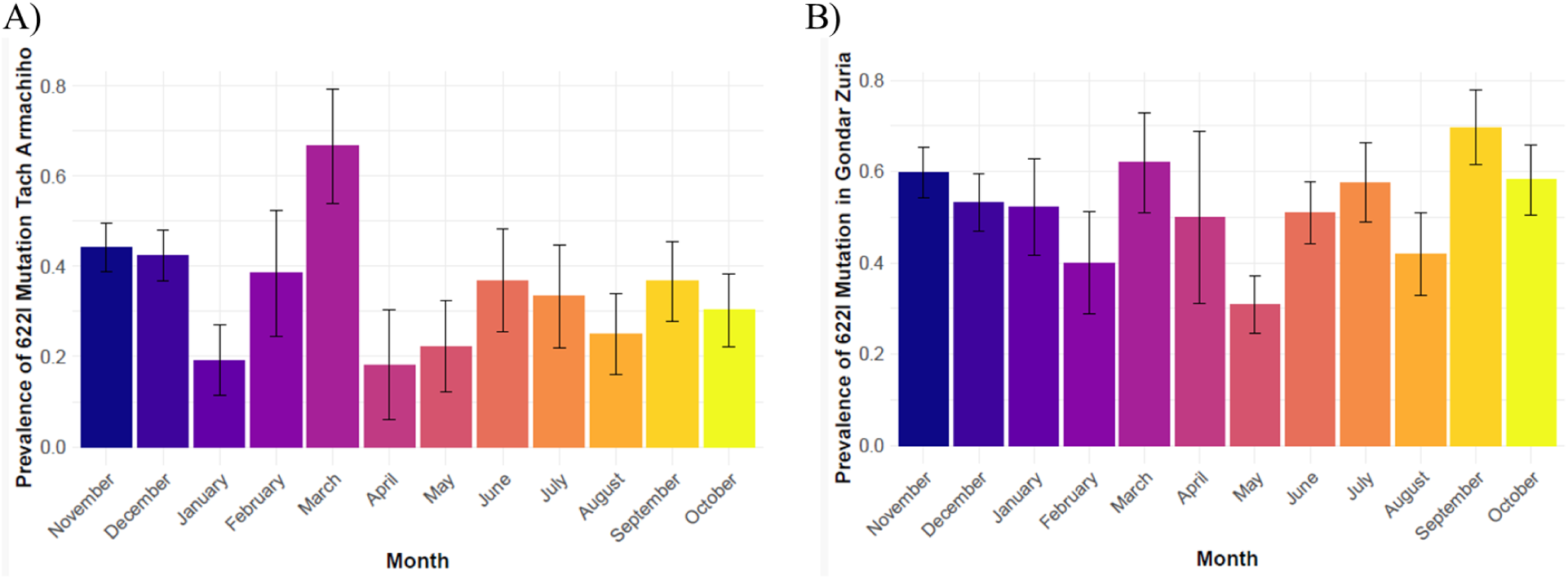
Panel **A** shows a monthly distribution of *k13* 622I prevalence in Tach Armachiho district. Panel **B** shows a monthly distribution of *k13* 622I prevalence in Gondar Zuria district. Error bars represent the 95% confidence intervals.

### Supplementary tables

To assess and view easily all supplementary tables compiled in one excel sheet.

**Supplementary Table 1.** Overall prevalence of key mutation among successfully genotyped drug resistance markers, N=number of samples successfully sequenced, n= number of samples with mutation and, %=prevalence of mutation.

**Supplementary Table 2.** Prevalence of key mutation among successfully genotyped drug resistance markers from each study district, n=number of samples successfully sequenced, and %=prevalence of mutation.

**Supplementary Table 3.** Prevalence of key mutation per Kebeles(Villages), n= number samples sequenced, and %=prevalence of key mutation.

**Supplementary Table 4.** Overall prevalence of 622I mutation per month, n= number of carriers for mutation, N=denominator (number of samples successfully sequenced), and %= 622I prevalence.

**Supplementary Table 5.** Monthly prevalence of 622I mutation in Gondar Zuria district, n= number of carriers for mutation, N=denominator (number of samples successfully sequenced), and %= R622I prevalence.

**Supplementary Table 6:** Monthly prevalence of 622I mutation in Tach Armachiho district, n=number of carriers for mutation, N=denominator (number of samples successfully sequenced), and %= R622I Prevalence.

### Supplementary Data

**Supplementary Data 1.** Metadata and genotype file for all successfully sequenced samples for drug resistance MIP panel. Only successfully sequenced samples were used for drug resistance marker prevalence estimates. 0=Reference Call, 1=Alternative heterozygous call, 2=Alternative homozygous call, −1=Missing call. Note that the number of samples successfully genotyped varies per locus drug resistance MIP panel.

## References

Abamecha, Abdulhakim, Daniel Yilma, Wondimagegn Adissu, Delenasaw Yewhalaw, and Alemseged Abdissa. 2021. “Efficacy and Safety of Artemether-Lumefantrine for Treatment of Uncomplicated Plasmodium Falciparum Malaria in Ethiopia: A Systematic Review and Meta-Analysis.” Malaria Journal 20 (1): 213.

Alemayehu, Aberham Abere, Daniel Castañeda Mogollón, Sisay Getie Belay, Habtie Tesfa, Abu Naser Mohon, Nirujah Balasingam, Abebe Genetu Bayih, Shoaib Ashraf, and Dylan R. Pillai. 2025. “Expansion of the Plasmodium Falciparum Kelch13 R622I Mutation in Northwest Ethiopia.” Open Forum Infectious Diseases 12 (6): ofaf279.

Anderson, T. J., B. Haubold, J. T. Williams, J. G. Estrada-Franco, L. Richardson, R. Mollinedo, M. Bockarie, et al. 2000. “Microsatellite Markers Reveal a Spectrum of Population Structures in the Malaria Parasite Plasmodium Falciparum.” Molecular Biology and Evolution 17 (10): 1467–82.

Asih, Puji B. S., William O. Rogers, Agustina I. Susanti, Agus Rahmat, Ismail E. Rozi, Mariska A. Kusumaningtyas, Krisin, et al. 2009. “Seasonal Distribution of Anti-Malarial Drug Resistance Alleles on the Island of Sumba, Indonesia.” Malaria Journal 8 (1): 222.

Aydemir, Ozkan, Mark Janko, Nick J. Hathaway, Robert Verity, Melchior Kashamuka Mwandagalirwa, Antoinette K. Tshefu, Sofonias K. Tessema, et al. 2018. “Drug-Resistance and Population Structure of Plasmodium Falciparum across the Democratic Republic of Congo Using High-Throughput Molecular Inversion Probes.” The Journal of Infectious Diseases 218 (6): 946–55.

Bayih, Abebe Genetu, Gebeyaw Getnet, Abebe Alemu, Sisay Getie, Abu Naser Mohon, and Dylan R. Pillai. 2016. “A Unique Plasmodium Falciparum K13 Gene Mutation in Northwest Ethiopia.” The American Journal of Tropical Medicine and Hygiene 94 (1): 132–35.

Bosman, Andrea, and Kamini N. Mendis. 2007. A Major Transition in Malaria Treatment: The Adoption and Deployment of Artemisinin-Based Combination Therapies. American Society of Tropical Medicine and Hygiene.

Brhane, Bokretsion G., Abebe A. Fola, Helen Nigussie, Alec Leonetti, Moges Kassa, Henok Hailgiorgis, Yonas Wuletaw, et al. 2024. “Rising Prevalence of Plasmodium Falciparum Artemisinin Resistance Mutations in Ethiopia.” Infectious Diseases (except HIV/AIDS). medRxiv. https://www.medrxiv.org/content/10.1101/2024.09.11.24313421v1.

Bugssa, Gessessew, and Kiros Tedla. 2020. “Feasibility of Malaria Elimination in Ethiopia.” Ethiopian Journal of Health Sciences 30 (4): 607–14.

Camponovo, Flavia, Caroline O. Buckee, and Aimee R. Taylor. 2023. “Measurably Recombining Malaria Parasites.” Trends in Parasitology 39 (1): 17–25.

Chang, Hsiao-Han, Colin J. Worby, Adoke Yeka, Joaniter Nankabirwa, Moses R. Kamya, Sarah G. Staedke, Grant Dorsey, et al. 2017. “THE REAL McCOIL: A Method for the Concurrent Estimation of the Complexity of Infection and SNP Allele Frequency for Malaria Parasites.” PLoS Computational Biology 13 (1): e1005348.

Connelly, Sean V., Julia G. Muller, Mohamed Ali, Billy E. Ngasala, Wahida Hassan, Bakari Mohamed, Kyaw L. Thwai, et al. 2025. “Artemisinin Partial Resistance Mutations in Zanzibar and Tanzania Suggest Regional Spread and African Origins, 2023.” Infectious Diseases (except HIV/AIDS). medRxiv. https://www.medrxiv.org/content/10.1101/2025.03.22.25323829v1.

Conrad, Melissa D., Victor Asua, Shreeya Garg, David Giesbrecht, Karamoko Niaré, Sawyer Smith, Jane F. Namuganga, et al. 2023. “Evolution of Partial Resistance to Artemisinins in Malaria Parasites in Uganda.” The New England Journal of Medicine 389 (8): 722–32.

Conway, Jake R., Alexander Lex, and Nils Gehlenborg. 2017. “UpSetR: An R Package for the Visualization of Intersecting Sets and Their Properties.” Bioinformatics 33 (18): 2938–40.

Daka, Demeke, Daniel Woldeyes, Lemu Golassa, Gezahegn Solomon Alemayehu, Zerihun Zewde, Girum Tamiru, Tadesse Misganaw, Fekadu Massebo, and Biniam Wondale. 2024. “Therapeutic Efficacy of Artemether-Lumefantrine in the Treatment of Uncomplicated Plasmodium Falciparum Malaria in Arba Minch Zuria District, Gamo Zone, Southwest Ethiopia.” Malaria Journal 23 (1): 282.

Deressa, Tekalign, Mengistu Endris Seid, Wubet Birhan, Yetemwork Aleka, and Biniam Mathewos Tebeje. 2017. “In Vivo Efficacy of Artemether-Lumefantrine against Uncomplicated Plasmodium Falciparum Malaria in Dembia District, Northwest Ethiopia.” Therapeutics and Clinical Risk Management 13 (February):201–6.

Dondorp, Arjen M., François Nosten, Poravuth Yi, Debashish Das, Aung Phae Phyo, Joel Tarning, Khin Maung Lwin, et al. 2009. “Artemisinin Resistance in Plasmodium Falciparum Malaria.” The New England Journal of Medicine 361 (5): 455–67.

Eloff, Lydia, Andrés Aranda-Díaz, Isobel Routledge, Amy Wesolowski, Mukosha Chisenga, Brighton Mangena, John Chimumbwa, et al. 2025. “High Prevalence of Molecular Markers Associated with Artemisinin, Sulphadoxine and Pyrimethamine Resistance in Northern Namibia.” Infectious Diseases (except HIV/AIDS). medRxiv. https://www.medrxiv.org/content/10.1101/2025.01.09.25320247v1.

Feleke, Sindew M., Emily N. Reichert, Hussein Mohammed, Bokretsion G. Brhane, Kalkidan Mekete, Hassen Mamo, Beyene Petros, et al. 2021. “Plasmodium Falciparum Is Evolving to Escape Malaria Rapid Diagnostic Tests in Ethiopia.” Nature Microbiology 6 (10): 1289–99.

Fola, Abebe A., Sindew M. Feleke, Hussein Mohammed, Bokretsion G. Brhane, Christopher M. Hennelly, Ashenafi Assefa, Rebecca M. Crudal, et al. 2023. “Plasmodium Falciparum Resistant to Artemisinin and Diagnostics Have Emerged in Ethiopia.” Nature Microbiology 8 (10): 1911–19.

Funk, Chris, Pete Peterson, Martin Landsfeld, Diego Pedreros, James Verdin, Shraddhanand Shukla, Gregory Husak, et al. 2015. “The Climate Hazards Infrared Precipitation with Stations--a New Environmental Record for Monitoring Extremes.” Scientific Data 2 (1): 150066.

Garrison, Erik, and Gabor Marth. 2012. “Haplotype-Based Variant Detection from Short-Read Sequencing.” arXiv [q-bio.GN]. arXiv. http://arxiv.org/abs/1207.3907.

Gesesew, Hailay, Kiros Berhane, Elias S. Siraj, Dawd Siraj, Mulugeta Gebregziabher, Yemane Gebremariam Gebre, Samuel Aregay Gebreslassie, et al. 2021. “The Impact of War on the Health System of the Tigray Region in Ethiopia: An Assessment.” BMJ Global Health 6 (11): e007328.

Grossman, Tamar, Julia Vainer, Yael Paran, Liora Studentsky, Uri Manor, Ron Dzikowski, and Eli Schwartz. 2023. “Emergence of Artemisinin-Based Combination Treatment Failure in Patients Returning from Sub-Saharan Africa with P. Falciparum Malaria.” Journal of Travel Medicine 30 (8): taad114.

Haile, Mebrahtom, Hailemariam Lemma, and Yemane Weldu. 2017. “Population Movement as a Risk Factor for Malaria Infection in High-Altitude Villages of Tahtay-Maychew District, Tigray, Northern Ethiopia: A Case-Control Study.” The American Journal of Tropical Medicine and Hygiene 97 (3): 726–32.

Halsey, Eric S., and Mateusz M. Plucinski. 2023. “Out of Africa: Increasing Reports of Artemether-Lumefantrine Treatment Failures of Uncomplicated Plasmodium Falciparum Infection.” Journal of Travel Medicine 30 (8). 10.1093/jtm/taad159.

Herman, Camelia, Curtis S. Huber, Sophie Jones, Laura Steinhardt, Mateusz M. Plucinski, Jean F. Lemoine, Michelle Chang, John W. Barnwell, Venkatachalam Udhayakumar, and Eric Rogier. 2019. “Multiplex Malaria Antigen Detection by Bead-Based Assay and Molecular Confirmation by PCR Shows No Evidence of Pfhrp2 and Pfhrp3 Deletion in Haiti.” Malaria Journal 18 (1): 380.

Ishengoma, Deus S., Celine I. Mandara, Catherine Bakari, Abebe A. Fola, Rashid A. Madebe, Misago D. Seth, Filbert Francis, et al. 2024. “Evidence of Artemisinin Partial Resistance in Northwestern Tanzania: Clinical and Molecular Markers of Resistance.” *The Lancet Infectious Diseases*, August. 10.1016/S1473-3099(24)00362-1.

Kamaliddin, Claire, Jack Burke-Gaffney, Shoaib Ashraf, Daniel Castañeda-Mogollón, Aderaw Adamu, Bacha Mekonen Tefa, Ayesha Wijesinghe, Enaara Pussegoda, Sindew Mekasha Feleke, and Dylan R. Pillai. 2024. “A Countrywide Survey of hrp2/3 Deletions and kelch13 Mutations Co-Occurrence in Ethiopia.” The Journal of Infectious Diseases 230 (6): e1394–1401.

Kokwaro, Gilbert, Leah Mwai, and Alexis Nzila. 2007. “Artemether/lumefantrine in the Treatment of Uncomplicated Falciparum Malaria.” Expert Opinion on Pharmacotherapy 8 (1): 75–94.

Kunte, Renuka, and Rajesh Kunwar. 2011. “WHO Guidelines for the Treatment of Malaria.” Armed Forces Medical Journal, India 67 (4): 376.

Lo, Eugenia, Elizabeth Hemming-Schroeder, Delenasaw Yewhalaw, Jennifer Nguyen, Estifanos Kebede, Endalew Zemene, Sisay Getachew, et al. 2017. “Transmission Dynamics of Co-Endemic Plasmodium Vivax and P. Falciparum in Ethiopia and Prevalence of Antimalarial Resistant Genotypes.” PLoS Neglected Tropical Diseases 11 (7): e0005806.

MalariaGEN, Muzamil Mahdi Abdel Hamid, Mohamed Hassan Abdelraheem, Desmond Omane Acheampong, Ambroise Ahouidi, Mozam Ali, Jacob Almagro-Garcia, et al. 2023. “Pf7: An Open Dataset of Plasmodium Falciparum Genome Variation in 20,000 Worldwide Samples.” Wellcome Open Research 8 (January):22.

Maltha, J., P. Gillet, and J. Jacobs. 2013. “Malaria Rapid Diagnostic Tests in Travel Medicine.” Clinical Microbiology and Infection: The Official Publication of the European Society of Clinical Microbiology and Infectious Diseases 19 (5): 408–15.

March, 1. n.d. “Ethiopia - Situation Report, 1 Mar 2024.” Accessed June 11, 2025. https://www.unocha.org/publications/report/ethiopia/ethiopia-situation-report-1-mar-2024.

Martin, Anne C., Jacob M. Sadler, Alfred Simkin, Michael Musonda, Ben Katowa, Japhet Matoba, Jessica Schue, et al. 2025. “Emergence and Rising Prevalence of Artemisinin Partial Resistance Marker Kelch13 P441L in a Low Malaria Transmission Setting in Southern Zambia.” Infectious Diseases (except HIV/AIDS). medRxiv. https://www.medrxiv.org/content/10.1101/2025.01.02.24319706v1.full.pdf.

Meier-Scherling, Cecile P. G., Oliver J. Watson, Victor Asua, Isaac Ghinai, Thomas Katairo, Shreeya Garg, Melissa Conrad, Philip J. Rosenthal, Lucy C. Okell, and Jeffrey A. Bailey. 2024. “Selection of Artemisinin Partial Resistance Kelch13 Mutations in Uganda in 2016-22 Was at a Rate Comparable to That Seen Previously in South-East Asia.” Infectious Diseases (except HIV/AIDS). medRxiv. https://www.medrxiv.org/content/10.1101/2024.02.03.24302209v1.full.

Mertens, Jonas E. 2024. “A History of Malaria and Conflict.” Parasitology Research 123 (3): 165.

Mihreteab, Selam, Lucien Platon, Araia Berhane, Barbara H. Stokes, Marian Warsame, Pascal Campagne, Alexis Criscuolo, et al. 2023. “Increasing Prevalence of Artemisinin-Resistant HRP2-Negative Malaria in Eritrea.” The New England Journal of Medicine 389 (13): 1191–1202.

Millet, Julie, Sandrine Alibert, Marylin Torrentino-Madamet, Christophe Rogier, Christiane Santelli-Rouvier, Patricia Bigot, Joel Mosnier, et al. 2004. “Polymorphism in Plasmodium Falciparum Drug Transporter Proteins and Reversal of in Vitro Chloroquine Resistance by a 9,10-Dihydroethanoanthracene Derivative.” Antimicrobial Agents and Chemotherapy 48 (12): 4869–72.

Niaré, Karamoko, Rebecca Crudale, Abebe A. Fola, Neeva Wernsman Young, Victor Asua, Melissa Conrad, Pierre Gashema, et al. 2025. “Highly Multiplex Molecular Inversion Probe Panel in Plasmodium Falciparum Targeting Common SNPs Approximates Whole Genome Sequencing Assessments for Selection and Relatedness.” medRxiv. 10.1101/2025.03.07.25323597.

Noedl, Harald, Youry Se, Kurt Schaecher, Bryan L. Smith, Duong Socheat, Mark M. Fukuda, and Artemisinin Resistance in Cambodia 1 (ARC1) Study Consortium. 2008. “Evidence of Artemisinin-Resistant Malaria in Western Cambodia.” The New England Journal of Medicine 359 (24): 2619–20.

Nosten, François, and Nicholas J. White. 2007. Artemisinin-Based Combination Treatment of Falciparum Malaria. American Society of Tropical Medicine and Hygiene.

Nzila, Alexis, John Okombo, Eric Ohuma, and Assad Al-Thukair. 2012. “Update on the in Vivo Tolerance and in Vitro Reduced Susceptibility to the Antimalarial Lumefantrine.” The Journal of Antimicrobial Chemotherapy 67 (10): 2309–15.

Ord, Rosalynn, Neal Alexander, Sam Dunyo, Rachel Hallett, Musa Jawara, Geoffrey Targett, Christopher J. Drakeley, and Colin J. Sutherland. 2007. “Seasonal Carriage of Pfcrt and pfmdr1 Alleles in Gambian Plasmodium Falciparum Imply Reduced Fitness of Chloroquine-Resistant Parasites.” The Journal of Infectious Diseases 196 (11): 1613–19.

Parobek, Christian M., Jonathan B. Parr, Nicholas F. Brazeau, Chanthap Lon, Suwanna Chaorattanakawee, Panita Gosi, Eric J. Barnett, et al. 2017. “Partner-Drug Resistance and Population Substructuring of Artemisinin-Resistant Plasmodium Falciparum in Cambodia.” Genome Biology and Evolution 9 (6): 1673–86.

Rogier, Eric, Jessica N. McCaffery, Doug Nace, Samaly Souza Svigel, Ashenafi Assefa, Jimee Hwang, Simon Kariuki, et al. 2022. “Plasmodium Falciparum pfhrp2 and pfhrp3 Gene Deletions from Persons with Symptomatic Malaria Infection in Ethiopia, Kenya, Madagascar, and Rwanda.” Emerging Infectious Diseases 28 (3): 608–16.

Schaffner, Stephen F., Aimee R. Taylor, Wesley Wong, Dyann F. Wirth, and Daniel E. Neafsey. 2018. “hmmIBD: Software to Infer Pairwise Identity by Descent between Haploid Genotypes.” Malaria Journal 17 (1): 196.

Schalkwyk, Donelly A. van, Sade Pratt, Debbie Nolder, Lindsay B. Stewart, Helen Liddy, Julian Muwanguzi-Karugaba, Khalid B. Beshir, et al. 2024. “Treatment Failure in a UK Malaria Patient Harboring Genetically Variant Plasmodium Falciparum from Uganda with Reduced in Vitro Susceptibility to Artemisinin and Lumefantrine.” Clinical Infectious Diseases: An Official Publication of the Infectious Diseases Society of America 78 (2): 445–52.

Slater, Hannah C., Jamie T. Griffin, Azra C. Ghani, and Lucy C. Okell. 2016. “Assessing the Potential Impact of Artemisinin and Partner Drug Resistance in Sub-Saharan Africa.” Malaria Journal 15 (1): 10.

Taffese, Hiwot S., Elizabeth Hemming-Schroeder, Cristian Koepfli, Gezahegn Tesfaye, Ming-Chieh Lee, James Kazura, Gui-Yun Yan, and Guo-Fa Zhou. 2018. “Malaria Epidemiology and Interventions in Ethiopia from 2001 to 2016.” Infectious Diseases of Poverty 7 (1): 103.

Teyssier, Noam B., Anna Chen, Elias M. Duarte, Rene Sit, Bryan Greenhouse, and Sofonias K. Tessema. 2021. “Optimization of Whole-Genome Sequencing of Plasmodium Falciparum from Low-Density Dried Blood Spot Samples.” Malaria Journal 20 (1): 116.

Uwimana, Aline, Eric Legrand, Barbara H. Stokes, Jean-Louis Mangala Ndikumana, Marian Warsame, Noella Umulisa, Daniel Ngamije, et al. 2020. “Emergence and Clonal Expansion of in Vitro Artemisinin-Resistant Plasmodium Falciparum kelch13 R561H Mutant Parasites in Rwanda.” Nature Medicine 26 (10): 1602–8.

Verity, Robert, Ozkan Aydemir, Nicholas F. Brazeau, Oliver J. Watson, Nicholas J. Hathaway, Melchior Kashamuka Mwandagalirwa, Patrick W. Marsh, et al. 2019. “The Impact of Antimalarial Resistance on the Genetic Structure of *Plasmodium Falciparum* in the DRC.” bioRxiv. 10.1101/656561.

Wernsman Young, Neeva, Pierre Gashema, David Giesbrecht, Tharcisse Munyaneza, Felicien Maisha, Fred Mwebembezi, Rule Budodo, et al. 2025. “High Frequency of Artemisinin Partial Resistance Mutations in the Great Lakes Region Revealed through Rapid Pooled Deep Sequencing.” The Journal of Infectious Diseases 231 (1): 269–80.

Yu, Qiwei, Ying Qu, Liqiang Zhang, Xin Yao, Jing Yang, Siyuan Chen, Hui Liu, et al. 2024. “Spatial Spillovers of Violent Conflict Amplify the Impacts of Climate Variability on Malaria Risk in Sub-Saharan Africa.” Proceedings of the National Academy of Sciences of the United States of America 121 (15): e2309087121.

Zheng, Zhenxian, Shumin Li, Junhao Su, Amy Wing-Sze Leung, Tak-Wah Lam, and Ruibang Luo. 2022. “Symphonizing Pileup and Full-Alignment for Deep Learning-Based Long-Read Variant Calling.” Nature Computational Science 2 (12): 797–803.

